# *APOE* missense variant R145C is associated with increased Alzheimer’s disease risk in African ancestry individuals with the *APOE ε3/ε4* genotype

**DOI:** 10.1101/2021.10.20.21265141

**Authors:** Yann Le Guen, Michael E. Belloy, Sarah J. Eger, Annabel Chen, Gabriel Kennedy, Timothy A. Thornton, Lindsay A. Farrer, Valerio Napolioni, Zihuai He, Michael D. Greicius

**Affiliations:** Department of Neurology and Neurological Sciences, Stanford University, Stanford, CA, 94304, USA; Institut du Cerveau - Paris Brain Institute - ICM, Paris, 75013, France; Department of Biostatistics, University of Washington, Seattle, WA, 98185, USA; Department of Medicine (Biomedical Genetics), Boston University School of Medicine, Boston, MA 02118, USA; School of Biosciences and Veterinary Medicine, University of Camerino, Camerino, 62032, Italy; Quantitative Sciences Unit, Department of Medicine, Stanford University, Stanford, CA, 94304, USA

**Author notes:** **Corresponding Author** Yann Le Guen, Department of Neurology and Neurological Sciences – Greicius lab, Stanford University, 290 Jane Stanford Way, E265, CA 94305-5090.

## Abstract

**BACKGROUND:** The *APOE* gene has two common missense variants that greatly impact the risk of late-onset Alzheimer’s disease (AD). Here we examined the risk of a third *APOE* missense variant, R145C, that is rare in European-Americans but present in 4% of African-Americans and always in phase with *APOE ε3*.

**METHODS:** In this study, we included 11,790 individuals of African and Admixed-African ancestry (4,089 cases and 7,701 controls). The discovery sample was composed of next generation sequencing data (2,888 cases and 4,957 controls), and the replication was composed of microarray data imputed on the TOPMed reference panel (1,201 cases and 2,744 contols). To assess the effect of R145C independently of the *ε2* and *ε4* alleles, we performed stratified analyses in *ε2/ε3, ε3/ε3*, and *ε3/ε4* subjects. In primary analyses, the AD risk associated with R145C was estimated using a linear mixed model regression on case-control diagnosis. In secondary analyses, we estimated the influence of R145C on age-at-onset using linear-mixed-model regression, and risk of conversion to AD using competing risk regression.

**RESULTS:** In *ε3/ε4-*stratified meta-analyses, R145C carriers had an almost three-fold increased risk compared to non-carriers (odds ratio, 2.75; 95% confidence interval [CI], 1.84 to 4.11; P = 8.3×10^−7^) and had a reported AD age-at-onset almost 6 years younger (β, -5.72; 95% CI, 7.87 to -3.56; P = 2.0×10^−7^). Competing risk regression showed that the cumulative incidence of AD grows faster with age in R145C carriers compared to non-carriers (hazard ratio, 2.42, 95% CI, 1.81 to 3.25; P = 3.7×10^−9^).

**CONCLUSION:** The R145C variant is a potent risk factor for AD among African ancestry individuals with the *ε3/ε4* genotype. Our findings should enhance AD risk prediction in African ancestry individuals and help elucidate the mechanisms linking the apoE protein to AD pathogenesis. The findings also add to the growing body of evidence demonstrating the importance of including ancestrally-diverse populations in genetic studies.

---

The *APOE ε4* allele and advanced age are the two main risk factors for late-onset Alzheimer’s disease (AD). The AD risk associated with common *APOE* missense mutations has long been established and replicated in people of European ancestry. In particular, as compared to the reference allele *ε3, ε4* increases the risk of AD^1^, and *ε2* is protective against AD^2^. The AD risk associated with these *APOE* alleles has been found to vary across ancestries^3^. Notably, the *ε4* associated risk is higher in Asians and lower in Africans, as compared to Europeans^3^. Several other missense variants have been identified on *APOE*. Some, such as Leu28Pro (L28P) and Val236Glu (V236E), are relatively common in individuals of European ancestry (0.1% to 1% of individuals). V236E has been associated with a reduced AD risk^4^, independently of the more common APOE missense variants. Another rare mutation, Arg136Ser (R136S), known as *APOE* Christchurch, may have a protective effect against early-onset AD in *PSEN1* mutation carriers^5^. The Christchurch finding should be considered preliminary, however, given that it is based on a single subject who was homozygous for R136S (no protective effect was seen in *PSEN1* mutation carriers with one copy of R136S)^5^. Other missense variants exist on *APOE* but have been understudied because they are present mainly or exclusively in individuals of African ancestry who have traditionally been underrepresented in studies of AD genetics. Here, we investigated whether *APOE* missense variants seen in African ancestry individuals affect AD risk. Apart from the variants defining the *ε2* and *ε4* alleles, the canonical *APOE* transcript only harbors two variants with minor allele count above 10 in our African ancestry discovery sample. One is common, Arg145Cys (R145C) with minor allele frequency (MAF) ≈ 2%, and one is relatively rare, Arg150His (R150H), with MAF ≈ 0.1% (**Table S1**). R145C and R150H are located at position rs769455 and rs376170967, respectively, both within the low-density lipoprotein receptor binding region of the apoE N-terminal domain, between the two variants, rs7412 and rs429358, used to determine the *ε2* and *ε4* alleles. The existing literature^6–8^ and the AD sequencing project (ADSP) data show that R145C is always observed in phase with *ε3* and R150H is always in phase with *ε2*. More than 4% of individuals of African ancestry carry R145C^6^ and it is linked to type III hyperlipoproteinemia (HLP)^6–8^. This variant is extremely rare in Europeans in gnomAD^9^ with a MAF ≈ 0.007%, which, combined with the fact that individuals of African ancestry are underrepresented in AD case/control datasets, explains why it has received little attention to date.

Two recent initiatives enabled us to assess the influence of these variants on AD risk. First, the ADSP which includes whole-exome sequencing (WES) and whole-genome sequencing (WGS) data, recently placed special emphasis on increasing genetic diversity, which expanded the number of non-European samples available^10^. Second, the TOPMed reference panel, which is both large and ancestrally diverse^11^, allowed us to impute African ancestry specific variants with high imputation quality.

Recent work has suggested that when examining variants at the *APOE* locus, *APOE*-stratified analyses may provide better statistical power than standard *APOE*-adjusted models^12,13^. In addition, *ε4* stratified analyses obviate concerns about appropriate modelling of the interaction term (e.g., linear vs multiplicative)^14^. Given that the two variants of interest here are at the *APOE* locus and that each is only seen on a unique *APOE* common haplotype, R145C on *ε3* and R150H on *ε2*, we used an *APOE*-stratified approach in our primary analysis and included standard *APOE*-adujsted models in secondary analyses.

## METHODS

### PARTICIPANTS AND SOURCES OF DATA

Participants or their caregivers provided written informed consent in the original studies. The current study protocol was granted an exemption by the Stanford University institutional review board because the analyses were carried out on deidentified, off-the-shelf data; therefore, additional informed consent was not required. Phenotypic information and genotypes were obtained from publicly released genome-wide association study datasets assembled by the Alzheimer’s Disease Genetics Consortium (ADGC) and derived from whole exome and whole genome sequence data generated by the Alzheimer Disease Sequencing Project (ADSP), with phenotype and genotype ascertainment described elsewhere^15–18,10^. The cohorts’ queried accession numbers, as well as the sequencing technology or single nucleotide polymorphism (SNP) genotyping platforms are described in **Tables S2 and S3**. Carriers of known pathogenic mutations on *APP, PSEN1, PSEN2*, and *MAPT* were excluded from our analysis. Discordant pathology cases, defined as any clinically diagnosed AD individual with Braak stage below III or neuritic plaque level below moderate, were excluded from our analysis.

### QUALITY CONTROL PROCEDURES

In each cohort-platform, variants were excluded based on genotyping rate (< 95%), MAF < 1%, and Hardy-Weinberg equilibrium in controls (p < 10^−6^) using PLINK v1.9^19^. gnomAD^9^ database-derived information was used to filter out SNPs that met one of the following exclusion criteria^20,21^: (i) located in a low complexity region, (ii) located within common structural variants (MAF > 1%), (iii) multiallelic SNPs with MAF > 1% for at least two alternate alleles, (iv) located within a common insertion/deletion, (v) having any flag different than PASS in gnomADv.3, (vi) having potential probe polymorphisms. The latter are defined as SNPs for which the probe may have variable affinity due to the presence of other SNP(s) within 20 bp and with MAF > 1%. Individuals with more than 5% genotype missingness were excluded. Duplicate individuals were identified with KING^22^ and their clinical, diagnostic and pathological data (including age-at-onset of cognitive symptoms, age-at-examination for clinical diagnosis, age-at-last exam, age-at-death), as well as sex, race, and *APOE* genotype were cross-referenced across cohorts. Duplicate entries with irreconcilable phenotype or discordant sex were flagged for exclusion. For individuals with duplicated genotype in sequencing and imputed data, the sequencing entry was used in the discovery set and the imputed entry was not included in the replication set. As some cohorts contributed to both the sequencing and genotyping platforms, some individuals in the discovery were related to individuals in the replication. Mega-analyses using linear mixed models that account for relatedness were run as sensitivity analyses (see Statistical analysis section).

### ANCESTRY DETERMINATION

For each cohort, we first determined the ancestry of each individual with SNPWeights v2^23^ using reference populations from the 1000 Genomes Consortium^24^. By applying an ancestry percentage cut-off > 75%, the samples were stratified into five super populations: South-Asians, East-Asians, Americans, Africans, and Europeans, and an Admixed group composed of individuals not passing the 75% cut-off in any single ancestry (**Table S4**) as implemented in our previous analyses on European individuals^20,21^. Since the *APOE* missense variants of interest (R145C and R150H) are extremely rare in non-African ancestry populations, we restricted our analysis to African and Admixed-African individuals. Admixed-African individuals included in the main analysis had at least 15% African ancestry, and we performed sensitivity analyses in increments of 30%, including Admixed-African individuals at 45% and 75% cutoffs. The latter corresponding to the super population threshold.

### IMPUTATION

Each cohort-genotyping platform was imputed on the TOPMed imputation server per ancestry group to obtain an imputation quality (R^2^) per ancestry group. For rs769455, R^2^ was ≤ 0.3 in all European ancestry cohorts, but was > 0.95 in most cohort-genotyping platforms for African and Admixed groups. This observation is consistent with the MAF > 2% in African Americans^9^ and MAF ≈ 0.007% in Europeans. We retained individuals with R^2^ > 0.8 at rs769455 (**Table S5**). As there was no signal for rs376170967 in the discovery sample, we did not end up imputing it in the replication datasets.

### *APOE* GENOTYPE ASCERTAINMENT

We directed specific attention to the genotyping of the SNPs determining the main *APOE* genotype (rs429358 and rs7412), rs769455-T (*APOE*[R145C]), and rs376170967-A (*APOE*[R150H]). Details are provided in a Supplementary Note.

### DISCOVERY AND REPLICATION SAMPLES

Our discovery sample was composed of African and Admixed-African ancestry individuals from the ADSP WES and WGS, corresponding to 2,888 AD cases and 4,957 cognitively normal controls (**Table 1**). To build a replication sample for R145C, we queried for individuals of African and Admixed-African ancestry in all of the publicly available microarray genetic datasets that we had access to at the time of the study in July 2021 (**Table 1**). Replication was not attempted for R150H as this variant showed no effect in the discovery sample analysis. After quality control and duplicate removal, 1,201 AD cases and 2,744 controls remained in the replication sample. **Table S6** presents the demographics of the remaining AD cases and cognitively unimpaired controls.

**Table 1.** Demographics per *APOE* genotype. DX: diagnosis, CN: cognitively normal, AD: Alzheimer’s disease, N: number of individuals, %Females: percentage of female individuals, μ and σ: mean age and standard error.

| Sample | DX | N | *APOE* $\epsilon 2/\epsilon 2$ | | *APOE* $\epsilon 2/\epsilon 3$ | | *APOE* $\epsilon 3/\epsilon 3$ | | *APOE* $\epsilon 2/\epsilon 4$ | | *APOE* $\epsilon 3/\epsilon 4$ | | *APOE* $\epsilon 4/\epsilon 4$ | |
| --- | --- | --- | --- | --- | --- | --- | --- | --- | --- | --- | --- | --- | --- | --- |
| | | | N<br>(%Females) | Age<br>$\mu(\sigma)$ | N<br>(%Females) | Age<br>$\mu(\sigma)$ | N<br>(%Females) | Age<br>$\mu(\sigma)$ | N<br>(%Females) | Age<br>$\mu(\sigma)$ | N<br>(%Females) | Age<br>$\mu(\sigma)$ | N<br>(%Females) | Age<br>$\mu(\sigma)$ |
| Discovery | CN | 4957 | 41<br>(58.5%) | 77.6<br>(9.2) | 709<br>(69.7%) | 77.6<br>(8.6) | 2622<br>(72.7%) | 77.2<br>(8.3) | 179<br>(76.5%) | 75.2<br>(8.8) | 1288<br>(72.2%) | 75.8<br>(8.4) | 118<br>(67.8%) | 73.0<br>(7.8) |
|  | AD | 2888 | 11<br>(81.8%) | 81.3<br>(5.4) | 225<br>(71.1%) | 79.6<br>(8.7) | 1145<br>(68.0%) | 78.2<br>(8.5) | 111<br>(67.6%) | 77.7<br>(8.7) | 1093<br>(68.9%) | 75.2<br>(8.7) | 303<br>(68.6%) | 70.7<br>(8.2) |
| Replication | CN | 2744 | 26<br>(69.2%) | 82.2<br>(6.7) | 377<br>(67.4%) | 80.7<br>(7.6) | 1318<br>(69.0%) | 78.7<br>(9.5) | 120<br>(67.5%) | 79.4<br>(8.8) | 786<br>(67.3%) | 78.3<br>(9.4) | 117<br>(76.9%) | 75.8<br>(9.9) |
|  | AD | 1201 | 5<br>(60.0%) | 78.2<br>(8.0) | 76<br>(57.9%) | 77.4<br>(9.7) | 430<br>(69.5%) | 76.4<br>(9.7) | 42<br>(64.3%) | 77.3<br>(8.4) | 491<br>(72.7%) | 73.9<br>(9.9) | 157<br>(64.3%) | 71.4<br>(8.6) |

### STATISTICAL ANALYSES

All statistical analyses were performed in R (v4.0.2). In primary analyses, we estimated the AD risk associated with R145C and R150H using a linear mixed model regression on case-control diagnosis in each *APOE* stratum. In secondary analyses, we estimated the influence of R145C on age-at-onset (AAO) using linear mixed model regression on AAO in AD cases, and risk of conversion to AD using competing risk regression. Secondary analyses were not conducted for R150H as this variant showed no effect in the primary analysis. The case-control and age-at-onset analyses used linear mixed model regression available through the *GENESIS* package (v3.12)^25^. Multivariate competing risk regression and cumulative incidence estimation were implemented using the *cmprsk* package (v2.2)^26^. In this time-to-event analysis, failure events were defined as age-at-onset for cases (conversion to AD) and age-at-death for controls. Controls without reported death were right censored at age-at-last-visit. Left censoring was set at 50 years old, and younger individuals were excluded from the analysis. As seen on the cumulative incidence curves (**Figure 1**), only a small percentage of cases, mainly *APOE*ε4/ε4, had an age-at-onset between 50 and 60 years in agreement with the prevalence distribution of late-onset AD. All statistical analyses were adjusted for sex and four genetic principal components estimated with the *PC-Air* method^27^ implemented in *GENESIS*. Linear mixed model analyses were additionally covaried by a sparse genetic relationship matrix estimated with the *PC-Relate* method^28^ implemented in *GENESIS*. Case-control analyses were not adjusted for age given that controls were older than cases in some *APOE ε3* genotype strata (**Table 1**). Correcting for age when cases are younger than controls leads to the model incorrectly inferring the age effect on AD risk, resulting in statistical power loss^20^. We ran additional analyses adjusted for age and observed equivalent results (**Table S7**) and, as expected from the incorrect inference of the age effect in the *ε3/ε4* samples, the significance in this *APOE* genotype was slightly reduced. The discovery analyses were considered significant if they reached a Bonferroni-corrected p-value threshold of 0.0125 (≈ 0.05/4), which accounts for the analysis of R145C in three *APOE* strata (*ε2/ε3, ε3/ε3*, and *ε3/ε4)* and R150H in *ε2/ε3*. R150H was also seen in *ε2/ε4* but in only 3 carriers so this stratum was not considered for analysis. There were no R150H carriers in the *ε2/ε2* stratum. Only the R145C *ε3/ε4*-stratified analyses were significant in the discovery, thus replications were considered significant at p < 0.05 with a concordant direction of effect. Our data and the existing literature^6–8^ show that R145C is always observed on the same chromosome copy as *ε3* (**Table 2**), while R150H is always observed on the same chromosome copy as *ε2*^6^. Thus, to assess the effect of R145C independently of *ε2* and *ε4*, we performed stratified analyses on the three main *APOE* genotypes which include R145C carriers: *ε2/ε3, ε3/ε3*, and *ε3/ε4*, and similarly for R150H carriers in *ε2/ε3*.

**Figure 1.**
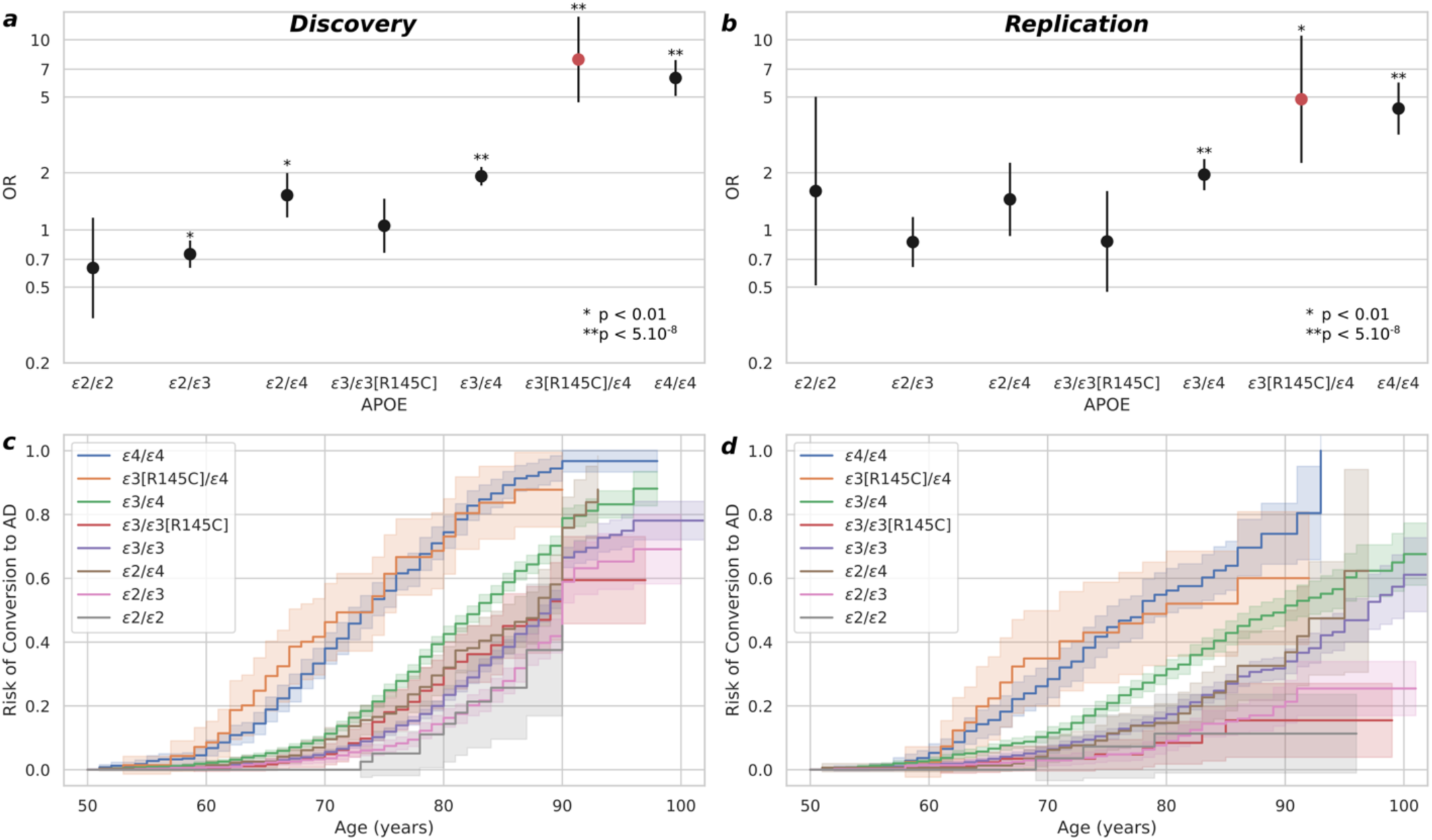
*APOE ε3[*R145C]/*ε4* individuals have an AD risk comparable to *APOE ε4/ε4* individuals. Alzheimer’s disease (AD) risk per *APOE* genotype assessed compared to the *APOE ε3/ε3* reference group (i.e., odds ratio (OR) *APOE ε3/ε3* equals 1) in (**a**) our discovery sample composed of next generation sequencing data from the ADSP dataset, and in (**b**) our replication composed of microarray data imputed on the TOPMed reference panel. AD cumulative incidence per *APOE* group, from the competing risk regression analysis accounting for the censored individuals (last visit or reported death), in the discovery (**c**) and replication (**d**).

**Table 2.** *APOE* R145C (rs769455) allelic breakdown by *APOE* genotype. Rs769455 alternate allele (T) is not observed in *APOE ε2/ε2, ε2/ε4, ε4/ε4*, and is only present in the homozygous state in *APOE ε3/ε3*, supporting the finding in sequencing databases that the alternate allele is always found in phase with *APOE ε3*. Note that rs769455 is located between rs7412 (99 bp apart) and rs429358 (39 bp apart) which define the *APOE* allele genotype. CN: cognitively normal, AD: Alzheimer’s disease, N: number of individuals.

| Sample | rs769455 | N total | *APOE* $\epsilon 2/\epsilon 2$ | | *APOE* $\epsilon 2/\epsilon 3$ | | *APOE* $\epsilon 3/\epsilon 3$ | | *APOE* $\epsilon 2/\epsilon 4$ | | *APOE* $\epsilon 3/\epsilon 4$ | | *APOE* $\epsilon 4/\epsilon 4$ | |
| --- | --- | --- | --- | --- | --- | --- | --- | --- | --- | --- | --- | --- | --- | --- |
|  |  |  | CN | AD | CN | AD | CN | AD | CN | AD | CN | AD | CN | AD |
| Discovery | C C | 7561 | 41 | 11 | 691 | 221 | 2490 | 1086 | 179 | 111 | 1269 | 1041 | 118 | 303 |
|  | C T | 279 | 0 | 0 | 18 | 4 | 129 | 57 | 0 | 0 | 19 | 52 | 0 | 0 |
|  | T T | 5 | 0 | 0 | 0 | 0 | 3 | 2 | 0 | 0 | 0 | 0 | 0 | 0 |
| Replication | C C | 3793 | 26 | 5 | 366 | 75 | 1236 | 416 | 120 | 42 | 765 | 468 | 117 | 157 |
|  | C T | 148 | 0 | 0 | 11 | 1 | 78 | 14 | 0 | 0 | 21 | 23 | 0 | 0 |
|  | T T | 4 | 0 | 0 | 0 | 0 | 4 | 0 | 0 | 0 | 0 | 0 | 0 | 0 |

To compare of our study with previous genome-wide association in African ancestry individuals^10^, we tested the R145C association with AD diagnosis using a standard linear mixed mode regression model, adjusting for *ε2* and *ε4* dosages, considering all individuals in **Table 1**. Given our main finding, with the R145C effect restricted to *APOE* ε3/ε4 carriers, we also tested for an R145C\**ε4* interaction in this standard model. Figures were plotted using the *seaborn* package (v.0.11.1) in Python (v3.9.4).

### *APOE* HAPLOTYPE LOCAL ANCESTRY ESTIMATION

To estimate the local ancestry of the *APOE* haplotype we considered a region encompassing the *APOE* gene with a 200kb-flank upstream and downstream (coordinates in build hg38 chr19:44705791-45109393). We phased separately the whole sample of ADSP WES and ADSP WGS using *Eagle* v2.4.1^29^, without using an external reference panel. Publicly available sequencing reference panels are much smaller than these two datasets and the *Eagle*’s documentation suggests that using a reference panel in this scenario is unlikely to significantly increase phasing accuracy. To estimate local ancestry we used *RFMix* v.2^30^ with the 893 AFR individuals and 633 EUR individuals from the expanded high-coverage (30x) whole-genome sequencing from the 1000 Genome Project data^31^. In sensitivity analyses, we re-analyzed the discovery sample data solely including individuals with AFR local ancestry at both *APOE* haplotypes.

## RESULTS

The R150H variant (rs376170967-A) is relatively rare in our data, seen in only 15 *ε2/ε3* carriers, and was not associated with AD risk in our primary analysis (odds ratio [OR], 1.08; 95% confidence interval [CI], 0.33 to 3.55; P = 0.90) As such, it was not investigated further.

In the discovery sample, *ε3/ε4* individuals who also carry the R145C variant (rs769455-T) had a three-fold increased risk of AD compared to *ε3/ε4* individuals lacking the R145C variant (odds ratio [OR], 3.01; 95% confidence interval [CI], 1.87 to 4.8; P = 6.0×10^−6^),. This association was significant in the replication sample (OR, 2.20; 95% CI, 1.04 to 4.65; P = 0.04). Among *ε3*/*ε4* subjects, those carrying the R145C variant also had an age-at-AD-onset more than 5 years younger than non-carriers in the discovery sample (β, -5.87; 95% CI, -8.35 to -3.4; P = 3.4×10^−6^) and in the replication sample (β, -5.23; 95% CI, - 9.58 to -0.87; P = 0.02). The competing risk results emphasized that the cumulative incidence of AD in *ε3/ε4* persons grows faster with age in individuals with, compared to those without, the R145C variant in the discovery sample (hazard ratio [HR], 2.66; 95% CI 1.86 to 3.80; P = 8.5×10^−8^) and in the replication sample (HR, 2.00; 95% CI 1.19 to 3.25; P = 8.7×10^−3^). The R145C variant was not associated with AD risk in individuals with the ε2/ε3 or ε3/ε3 genotype (**Table 3**).

**Table 3.** R145C is associated with increased AD risk and with younger age-at-onset specifically in *APOE ε3/ε4* individuals. Since R145C is in phase with *APOE ε3*, stratified analyses were limited to *APOE ε2/ε3, APOE ε3/ε3*, and *APOE ε3/ε4* genotypes. Discovery sample is composed of next generation sequencing data, while replication sample included microarray data imputed on the TOPMed reference panel. *APOE ε3[R145C]/ε4* individuals have significantly higher AD risk, younger onset, and higher risk of conversion from healthy aging to AD than *APOE ε3/ε4* individuals. N: number of individuals, MAC: minor allele count, OR: odds ratio, β: parameter estimate in the regression, HR: hazard ratio, P: p-value.

|  | Sample | AD Case-Control Regression |  |  |  | AD Age-at-onset Regression |  |  |  | Competing Risk Regression |  |  |  |
| --- | --- | --- | --- | --- | --- | --- | --- | --- | --- | --- | --- | --- | --- |
| | | N | MAC | OR | P | N | MAC | $\beta$ | P | N | MAC | HR | P |
|  |  |  |  | [95% CI] |  |  |  | [95% CI] |  |  |  | [95% CI] |  |
| *APOE* $\epsilon 2/\epsilon 3$ | *Discovery* | 934 | 22 | 0.73<br>[0.26; 2.04] | 0.55 | 222 | 4 | -6.96<br>[-15.56; 1.64] | 0.11 | 918 | 22 | 1.14<br>[0.35; 3.69] | 0.82 |
|  | *Replication* | 453 | 12 | 0.78<br>[0.11; 5.35] | 0.80 | 53 | 1 | -18.42<br>[-39.23; 2.38] | 0.08 | 430 | 12 | 3.05<br>[0.33; 28.09] | 0.32 |
|  | **Meta-analysis** | 1387 | 34 | 0.74<br>[0.3; 1.84] | 0.52 | 275 | 5 | -8.63<br>[-16.58; -0.69] | 0.03 | 1348 | 34 | 1.42<br>[0.5; 3.99] | 0.51 |
| *APOE* $\epsilon 3/\epsilon 3$ | *Discovery* | 3767 | 196 | 1.06<br>[0.78; 1.46] | 0.71 | 1108 | 58 | -1.68<br>[-3.87; 0.5] | 0.13 | 3646 | 187 | 1.09<br>[0.82; 1.45] | 0.57 |
|  | *Replication* | 1748 | 100 | 0.85<br>[0.48; 1.53] | 0.60 | 347 | 8 | -1.36<br>[-8.29; 5.58] | 0.70 | 1656 | 94 | 0.80<br>[0.41; 1.57] | 0.51 |
|  | **Meta-analysis** | 5515 | 296 | 1.01<br>[0.77; 1.34] | 0.94 | 1455 | 66 | -1.65<br>[-3.74; 0.43] | 0.12 | 5302 | 281 | 1.04<br>[0.8; 1.35] | 0.79 |
| *APOE* $\epsilon 3/\epsilon 4$ | *Discovery* | 2381 | 71 | 3.01<br>[1.87; 4.85] | **$6.0 \times 10^{-6}$** | 1063 | 51 | -5.87<br>[-8.35; -3.4] | **$3.4 \times 10^{-6}$** | 2315 | 70 | 2.66<br>[1.86; 3.8] | **$8.5 \times 10^{-8}$** |
|  | *Replication* | 1277 | 44 | 2.20<br>[1.04; 4.65] | **0.04** | 421 | 21 | -5.23<br>[-9.58; -0.87] | **0.02** | 1195 | 42 | 2.00<br>[1.19; 3.35] | **$8.7 \times 10^{-3}$** |
|  | **Meta-analysis** | 3658 | 115 | 2.75<br>[1.84; 4.11] | **$8.3 \times 10^{-7}$** | 1484 | 72 | -5.72<br>[-7.87; -3.56] | **$2.0 \times 10^{-7}$** | 3510 | 112 | 2.42<br>[1.81; 3.25] | **$3.7 \times 10^{-9}$** |

Results of sensitivity analyses evaluating different African ancestry cutoffs are shown in **Table S8**. Briefly, the results remained unchanged when selecting Admixed ancestry individuals with at least 45% African ancestry, or when restricting the analysis to African ancestry individuals (75% cutoff). We note that the OR increases with the African ancestry cutoff. For example, using an ancestry cutoff at 75% in the discovery cohort yielded an odds ratio of 3.40 (95% CI, 1.95 to 5.90; P = 1.5×10^−5^), as compared to an odds ratio of 3.01 using a cutoff of 15%. The results remain significant independent of this cutoff. Additionally, restricting our *ε3/ε4*-stratified analyses to individuals with estimated local ancestry AFR on both *APOE* haplotypes led to similar effect sizes (OR, 3.10; 95% CI, 1.68 to 5.70; P = 2.8×10^−4^) with lower significance due to substantially reduced sample sizes (**Table S9**). These results suggest that our analyses are not confounded by differences in local ancestry and that R145C is the causal variant. To account for the related individuals across the discovery and replication, we re-ran these analyses using a mega-analysis design, merging the discovery and replication samples and using linear mixed models (that account for relatedness) to test the association with AD diagnosis and age-at-onset. Our results remain unchanged and even slightly improved compared to the meta-analysis. Notably in the *ε3/ε4* group the significance of associations with AD risk (OR, 2.93; 95% CI, 1.99 to 4.31; P = 4.8×10^−8^), and age-at-onset (β, -5.86; 95% CI, -8.05 to -3.66; P = 1.7×10^−7^) both increased (**Table S10**).

To compare our study with earlier genome-wide association studies we conducted a standard (non-stratified) mixed mode regression model, adjusting for ε2 and ε4 dosages. As expected, given the specifity of the main effect in subjects with the *APOE ε3/ε4* genotype, the odds ratio was smaller than in the *ε3/ε4*-stratified analysis (OR, 1.36; 95% CI, 1.08 to 1.69; P = 0.0075, **Table S11**). Of note, Kunkle et al.^10^ reported a similar odds ratio in their equivalent analysis (OR, 1.38; 95% CI, 0.98 to 1.94; P = 0.056) conducted on a slightly smaller subset of samples used here. Kunkle et al. ^10^ did not, however, test the interaction of R145C with *ε4* for association with AD status. We formally tested this interaction and found a significant association (OR, 2.66; 95% CI, 1.68 to 4.23; P = 3.4×10^−5^, **Table S12**), supporting the main finding of R145C being associated with increased risk only in *APOE ε3/ε4* subjects.

We estimated the odds per *APOE* genotype group, using *ε3*/*ε3* individuals, non-carriers of R145C as the reference (i.e., odds ratio of *APOEε3*/*ε3* individuals equals 1). Strikingly, the odds ratios for AD among *ε3/ε4* individuals carrying the R145C missense variant and among *ε4/ε4* homozygotes are very similar in both the discovery (**Figure 1a**) and replication (**Figure 1b**) datasets. Sensitivity analyses in the 45% and 75% ancestry cutoffs lead to the same conclusion (**Figure S1**). Competing risk analyses conducted within each *APOE* genotype group also suggest that *ε3*/*ε4* individuals with the R145C missense variant have a similar cumulative incidence distribution as *ε4/ε4* individuals, and significantly different distributions from *ε3/ε4* individuals without the R145C missense variant in both the discovery (**Figure 1c**) and replication analyses (**Figure 1d**). Sensitivity analyses attained equivalent results at African ancestry cutoffs of 45% and 75% (**Figure S2**), suggesting that the significance of our results is independent of the African ancestry cutoffs and not due to population stratification potentially driven by Admixed individuals.

## DISCUSSION

We have shown that the R145C missense variant more than doubles AD risk in African ancestry individuals with the common *ε3/ε4* genotype. This variant is found in roughly 4% of African-Americans. As the field begins to design and undertake clinical trials stratified by *APOE* genotype, or even targeting *ε4* carriers exclusively, it will be important to weigh the substantial increase in risk conveyed by R145C^32^. Similarly, in the age of direct-to-consumer genetic testing where, increasingly, patients come to physicians with their *APOE* genotype in hand, care providers will need to understand AD risk according to specific ancestral backgrounds in order to provide optimized counseling.

Regarding potential mechanisms driving this effect, the R145C mutation is located within apoE’s receptor-binding region at amino acid residues 136 to 150 in the N-terminal domain. Mutations in this region, including R145C, have been linked to autosomal dominant type III hyperlipoproteinemia (HLP)^6–8,33,34^. Studies of R145C have suggested numerous effects on the protein that could account for the increased risk of type III HLP and might relate to the increased AD risk shown here. In particular, R145C was shown to partially inhibit apoE’s binding to the VLDL receptor^35^. Additionally, when compared to the apoE-*ε3* protein, the apoE-*ε3[*R145C] isoform binds less avidly to heparan sulfate proteoglycans (HSPG)^36^. Several studies have identified a role of HSPG in the cellular uptake of both Aβ^37,38^ and tau^39,40^ and it has been shown that apoE can compete with Aβ at this receptor^37,38^.

While there are many mechanisms that might link R145C to AD pathogenesis, the results reported here may provide some additional guidance. One of the more striking findings of this study is that the increased risk associated with R145C was only seen on an *ε3/ε4* background. While the risk of *ε3[*R145C]/*ε4* individuals is, in fact, similar to that of *ε4/ε4* carriers (**Figure 1**), we did not detect any, even suggestive, signal for R145C in *ε3/ε3* individuals. Furthermore, although we were not adequately powered to estimate its effect in the homozygous state, in our data we had 9 R145C homozygotes and their distribution across diagnoses (2 cases and 7 controls) supports the main finding that this variant only increases risk when paired with *ε4* on the other chromosome (**Table 2**). The additional cysteine in R145C carriers raises the possibility that a novel disulfide bond within apoE-*ε3* could alter its conformation^41,42^. If, in the normal state, apoE-*ε3* is able to mitigate the increased risk associated with apoE-*ε4*, R145C-induced changes in apoE-*ε3* presumably prevent this mitigation. This would be consistent with our finding that R145C has no impact on AD risk in *ε3/ε3* homozygotes and with the fact that *ε3/ε4* individuals with R145C have a risk profile similar to *ε4/ε4* homozygotes since, in both the *ε4/ε4* and the *ε3[*R145C]/*ε4* genotypes, the apoE-*ε4* effect is unmitigated by the normal apoE-*ε3* protein.

These results will, hopefully, spur additional investigations into the impact of R145C on apoE-*ε3* function, elucidating, in turn, the role of *APOE* in AD pathogenesis. This study also emphasizes the importance of recent efforts to enroll more diverse populations in studies of complex genetic diseases. Such efforts will lead not only to better, ancestry-informed risk estimates for individuals, but will also allow us to discover ancestry-specific variants offering critical new insights into disease mechanisms and, ultimately, drug development.

## Data Availability

Data availability
Data used in preparation of this manuscript can be obtained upon application at:
- dbGaP (https://www.ncbi.nlm.nih.gov/gap/advanced_search/)
- NIAGADS and NIAGADS DSS (https://www.niagads.org/)
- LONI (https://ida.loni.usc.edu/)
- Synapse (https://adknowledgeportal.synapse.org/)
- RADC Rush (https://www.radc.rush.edu/)
- NACC (https://naccdata.org/)
Tables S2 and S3 provide the details of repositories and accession number per cohort-platform group.

https://www.niagads.org/

https://ida.loni.usc.edu/

https://adknowledgeportal.synapse.org/

https://www.radc.rush.edu/

https://naccdata.org/

## FUNDINGS AND ACKNOWLEDGMENTS

Supported by the National Institute of Health and National Institute of Aging grants AG060747 (MDG), AG066206 (ZH), AG066515 (ZH, MDG), 2R01-AG048927 (LAF), RF1-AG057519 (LAF), U19-AG068753 (LAF), U01-AG058654 (LAF), U01-AG062602 (LAF), the European Union’s Horizon 2020 research and innovation program under the Marie Skłodowska-Curie (grant agreement No. 890650, YLG), the Alzheimer’s Association (AARF-20-683984, MEB), and the Iqbal Farrukh and Asad Jamal Fund. Additional funders of individual investigators and institutions who contributed to data collection and genotyping are provided in the **Supplementary Appendix**.

## Data availability

Data used in preparation of this manuscript can be obtained upon application at:

- dbGaP (https://www.ncbi.nlm.nih.gov/gap/advanced_search/)
- NIAGADS and NIAGADS DSS (https://www.niagads.org/)
- LONI (https://ida.loni.usc.edu/)
- Synapse (https://adknowledgeportal.synapse.org/)
- RADC Rush (https://www.radc.rush.edu/)
- NACC (https://naccdata.org/)

**Tables S2 and S3** provide the details of repositories and accession number per cohort-platform group.

