## Supplementary Appendix for "*APOE* missense variant R145C is associated with increased Alzheimer’s disease risk in African ancestry individuals with the *APOE ε3/ε4* genotype"

**Corresponding Author**

### Table of Contents


#### Supplementary Acknowledgments……………………...3

#### Supplementary Note………………………….........13

*APOE* GENOTYPE ASCERTAINMENT…………………………….....13

#### Supplementary Figures………………………………16

Figure S1. *APOEε3/*R145C]/*ε4* individuals have an AD risk comparable to *APOEε4/ε4* individuals regardless of the African ancestry cutoff…………………………………16

Figure S2. The *APOEε3/*R145C]/*ε4* cumulative incidence closely mirrors the *APOEε4/ε4* cumulative incidence regardless of the African ancestry cutoff……………………..17

Figure S3. Flowchart describing the *APOE* alleles and R145C genotyping among African and Admixed-African individuals included in the discovery analysis……..18

#### Supplementary Tables…………………………..…..19

Table S1. Missense mutations on APOE canonical transcript ……………………………………19

Table S2. Cohorts overview…………………………………………………………………...….20

Table S3. Overview of Alzheimer's disease sequencing project studies……………………..21

Table S4. Demographics of all the queried cohorts………………………………………..…...22

Table S5. R145C per cohort, diagnosis and *APOE* genotypes…………………………..………23

Table S6. Demographics per cohort after ancestry selection, quality control and duplicates removal……..……………………………..……………………………………………………...24

Table S7. African ancestry cutoff sensitivity analyses adding age as a covariate………………25

Table S8. African ancestry cutoff sensitivity analyses……………………………………………..27

Table S9. African ancestry cutoff sensitivity analyses restricted to individuals with AFR local ancestry at both of their *APOE* haplotypes……………………………………………………....29

Table S10. Mega-analysis of the discovery and replication samples………………….…………30

Table S11. Primary and secondary analyses considering standard model (non-stratified by *APOE* genotype) and adjusting for ε2 and ε4 dosages. ………………………………………...….31

Table S12. Interaction between R145C*ε4 for association with AD risk, considering standard model (non-stratified by *APOE* genotype) and adjusting for ε2 and ε4 dosages………..…….31

Table S13. *APOE* variants genotype call in the data released by NIAGADS…………………..32

Table S14 African ancestry cutoff sensitivity analyses only including individuals directly genotyped in the ADSP WES data distributed by NIAGADS……………………………….…..33

#### References………………………………….....................34

### Additional Acknowledgments

Data for this study were prepared, archived, and distributed by the National Institute on Aging Alzheimer's Disease Data Storage Site (NIAGADS) at the University of Pennsylvania (U24-AG041689), funded by the National Institute on Aging.

**Acknowledgments for the use of ADSP WES and WGS data**

The Alzheimer's Disease Sequencing Project (ADSP) is comprised of two Alzheimer's Disease (AD) genetics consortia and three National Human Genome Research Institute (NHGRI) funded Large Scale Sequencing and Analysis Centers (LSAC). The two AD genetics consortia are the Alzheimer's Disease Genetics Consortium (ADGC) funded by NIA (U01 AG032984), and the Cohorts for Heart and Aging Research in Genomic Epidemiology (CHARGE) funded by NIA (R01 AG033193), the National Heart, Lung, and Blood Institute (NHLBI), other National Institute of Health (NIH) institutes and other foreign governmental and non-governmental organizations. The Discovery Phase analysis of sequence data is supported through UF1AG047133 (to Drs. Schellenberg, Farrer, Pericak-Vance, Mayeux, and Haines); U01AG049505 to Dr. Seshadri; U01AG049506 to Dr. Boerwinkle; U01AG049507 to Dr. Wijsman; and U01AG049508 to Dr. Goate and the Discovery Extension Phase analysis is supported through U01AG052411 to Dr. Goate, U01AG052410 to Dr. Pericak-Vance and U01 AG052409 to Drs. Seshadri and Fornage.

Sequencing for the Follow Up Study (FUS) is supported through U01AG057659 (to Drs. PericakVance, Mayeux, and Vardarajan) and U01AG062943 (to Drs. Pericak-Vance and Mayeux). Data generation and harmonization in the Follow-up Phase is supported by U54AG052427 (to Drs. Schellenberg and Wang). The FUS Phase analysis of sequence data is supported through U01AG058589 (to Drs. Destefano, Boerwinkle, De Jager, Fornage, Seshadri, and Wijsman), U01AG058654 (to Drs. Haines, Bush, Farrer, Martin, and Pericak-Vance), U01AG058635 (to Dr. Goate), RF1AG058066 (to Drs. Haines, Pericak-Vance, and Scott), RF1AG057519 (to Drs. Farrer and Jun), R01AG048927 (to Dr. Farrer), and RF1AG054074 (to Drs. Pericak-Vance and Beecham).

The ADGC cohorts include: Adult Changes in Thought (ACT) (UO1 AG006781, UO1 HG004610, UO1 HG006375, U01 HG008657), the Alzheimer's Disease Centers (ADC) ( P30 AG019610, P30 AG013846, P50 AG008702, P50 AG025688, P50 AG047266, P30 AG010133, P50 AG005146, P50 AG005134, P50 AG016574, P50 AG005138, P30 AG008051, P30 AG013854, P30 AG008017, P30 AG010161, P50 AG047366, P30 AG010129, P50 AG016573, P50 AG016570, P50 AG005131, P50 AG023501, P30 AG035982, P30 AG028383, P30 AG010124, P50 AG005133, P50 AG005142, P30 AG012300, P50 AG005136, P50 AG033514, P50 AG005681, and P50 AG047270), the Chicago Health and Aging Project (CHAP) (R01 AG11101, RC4 AG039085, K23 AG030944), Indianapolis Ibadan (R01 AG009956, P30 AG010133), the Memory and Aging Project (MAP) ( R01 AG17917), Mayo Clinic (MAYO) (R01 AG032990, U01 AG046139, R01 NS080820, RF1 AG051504, P50 AG016574), Mayo Parkinson's Disease controls (NS039764, NS071674, 5RC2HG005605), University of Miami (R01 AG027944, R01 AG028786, R01 AG019085, IIRG09133827, A2011048), the Multi-Institutional Research in Alzheimer's Genetic Epidemiology Study (MIRAGE) (R01 AG09029, R01 AG025259), the National Cell Repository for Alzheimer's Disease (NCRAD) (U24

AG21886), the National Institute on Aging Late Onset Alzheimer's Disease Family Study (NIA-LOAD) (R01 AG041797), the Religious Orders Study (ROS) (P30 AG10161, R01 AG15819), the Texas Alzheimer's Research and Care Consortium (TARCC) (funded by the Darrell K Royal Texas Alzheimer's Initiative), Vanderbilt University/Case Western Reserve University (VAN/CWRU) (R01 AG019757, R01 AG021547, R01 AG027944, R01 AG028786, P01 NS026630, and Alzheimer's Association), the Washington Heights-Inwood Columbia Aging Project (WHICAP) (RF1 AG054023), the University of Washington Families (VA Research Merit Grant, NIA: P50AG005136, R01AG041797, NINDS: R01NS069719), the Columbia University HispanicEstudio Familiar de Influencia Genetica de Alzheimer (EFIGA) (RF1 AG015473), the University of Toronto (UT) (funded by Wellcome Trust, Medical Research Council, Canadian Institutes of Health Research), and Genetic Differences (GD) (R01 AG007584). The CHARGE cohorts are supported in part by National Heart, Lung, and Blood Institute (NHLBI) infrastructure grant HL105756 (Psaty), RC2HL102419 (Boerwinkle) and the neurology working group is supported by the National Institute on Aging (NIA) R01 grant AG033193.

The CHARGE cohorts participating in the ADSP include the following: Austrian Stroke Prevention Study (ASPS), ASPS-Family study, and the Prospective Dementia Registry-Austria (ASPS/PRODEM-Aus), the Atherosclerosis Risk in Communities (ARIC) Study, the Cardiovascular Health Study (CHS), the Erasmus Rucphen Family Study (ERF), the Framingham Heart Study (FHS), and the Rotterdam Study (RS). ASPS is funded by the Austrian Science Fond (FWF) grant number P20545-P05 and P13180 and the Medical University of Graz. The ASPS-Fam is funded by the Austrian Science Fund (FWF) project I904),the EU Joint Programme - Neurodegenerative Disease Research (JPND) in frame of the BRIDGET project (Austria, Ministry of Science) and the Medical University of Graz and the Steiermärkische Krankenanstalten Gesellschaft. PRODEM-Austria is supported by the Austrian Research Promotion agency (FFG) (Project No. 827462) and by the Austrian National Bank (Anniversary Fund, project 15435. ARIC research is carried out as a collaborative study supported by NHLBI contracts (HHSN268201100005C, HHSN268201100006C, HHSN268201100007C, HHSN268201100008C, HHSN268201100009C, HHSN268201100010C, HHSN268201100011C, and HHSN268201100012C). Neurocognitive data in ARIC is collected by U01 2U01HL096812, 2U01HL096814, 2U01HL096899, 2U01HL096902, 2U01HL096917 from the NIH (NHLBI, NINDS, NIA and NIDCD), and with previous brain MRI examinations funded by R01-HL70825 from the NHLBI. CHS research was supported by contracts HHSN268201200036C, HHSN268200800007C, N01HC55222, N01HC85079, N01HC85080, N01HC85081, N01HC85082, N01HC85083, N01HC85086, and grants U01HL080295 and U01HL130114 from the NHLBI with additional contribution from the National Institute of Neurological Disorders and Stroke (NINDS). Additional support was provided by R01AG023629, R01AG15928, and R01AG20098 from the NIA. FHS research is supported by NHLBI contracts N01-HC-25195 and HHSN268201500001I. This study was also supported by additional grants from the NIA (R01s AG054076, AG049607 and AG033040 and NINDS (R01 NS017950). The ERF study as a part of EUROSPAN (European Special Populations Research Network) was supported by European Commission FP6 STRP grant number 018947 (LSHG-CT-2006-01947) and also received funding from the European Community's Seventh Framework Programme (FP7/2007-2013)/grant agreement HEALTH-F4- 2007-201413 by the European Commission under the programme "Quality of Life and Management of the Living Resources" of 5th Framework Programme (no. QLG2-CT-2002- 01254). High-throughput analysis of the ERF data was supported by a joint grant from the Netherlands Organization for Scientific Research and the Russian Foundation for Basic Research (NWO-RFBR 047.017.043). The Rotterdam Study is funded by Erasmus Medical Center and Erasmus University, Rotterdam, the Netherlands Organization for Health Research and Development (ZonMw), the Research Institute for Diseases in the Elderly (RIDE), the Ministry of Education, Culture and Science, the Ministry for Health, Welfare and Sports, the European Commission (DG XII), and the municipality of Rotterdam. Genetic data sets are also supported by the Netherlands Organization of Scientific Research NWO Investments (175.010.2005.011, 911-03-012), the Genetic Laboratory of the Department of Internal Medicine, Erasmus MC, the Research Institute for Diseases in the Elderly (014-93-015; RIDE2), and the Netherlands Genomics Initiative (NGI)/Netherlands Organization for Scientific Research (NWO) Netherlands Consortium for Healthy Aging (NCHA), project 050-060-810. All studies are grateful to their participants, faculty and staff. The content of these manuscripts is solely the responsibility of the authors and does not necessarily represent the official views of the National Institutes of Health or the U.S. Department of Health and Human Services.

The FUS cohorts include: the Alzheimer's Disease Centers (ADC) ( P30 AG019610, P30 AG013846, P50 AG008702, P50 AG025688, P50 AG047266, P30 AG010133, P50 AG005146, P50 AG005134, P50 AG016574, P50 AG005138, P30 AG008051, P30 AG013854, P30 AG008017, P30 AG010161, P50 AG047366, P30 AG010129, P50 AG016573, P50 AG016570, P50 AG005131, P50 AG023501, P30 AG035982, P30 AG028383, P30 AG010124, P50 AG005133, P50 AG005142, P30 AG012300, P50 AG005136, P50 AG033514, P50 AG005681, and P50 AG047270), Alzheimer's Disease Neuroimaging Initiative (ADNI) (U19AG024904), Amish Protective Variant Study (RF1AG058066), Cache County Study (R01AG11380, R01AG031272, R01AG21136, RF1AG054052), Case Western Reserve University Brain Bank (CWRUBB) (P50AG008012), Case Western Reserve University Rapid Decline (CWRURD) (RF1AG058267, NU38CK000480), CubanAmerican Alzheimer's Disease Initiative (CuAADI) (3U01AG052410), Estudio Familiar de Influencia Genetica en Alzheimer (EFIGA) (5R37AG015473, RF1AG015473, R56AG051876), Genetic and Environmental Risk Factors for Alzheimer Disease Among African Americans Study (GenerAAtions) (2R01AG09029, R01AG025259, 2R01AG048927), Gwangju Alzheimer and Related Dementias Study (GARD) (U01AG062602), Hussman Institute for Human Genomics Brain Bank (HIHGBB) (R01AG027944, Alzheimer's Association "Identification of Rare Variants in Alzheimer Disease"), Ibadan Study of Aging (IBADAN) (5R01AG009956), Mexican Health and Aging Study (MHAS) (R01AG018016), Multi-Institutional Research in Alzheimer's Genetic Epidemiology (MIRAGE) (2R01AG09029, R01AG025259, 2R01AG048927), Northern Manhattan Study (NOMAS) (R01NS29993), Peru Alzheimer's Disease Initiative (PeADI) (RF1AG054074), Puerto Rican 1066 (PR1066) (Wellcome Trust (GR066133/GR080002), European Research Council (340755)), Puerto Rican Alzheimer Disease Initiative (PRADI) (RF1AG054074), Reasons for Geographic and Racial Differences in Stroke (REGARDS) (U01NS041588), Research in African American Alzheimer Disease Initiative (REAAADI) (U01AG052410), Rush Alzheimer's Disease Center (ROSMAP) (P30AG10161, R01AG15819, R01AG17919), University of Miami Brain Endowment Bank (MBB), and University of Miami/Case Western/North Carolina A&T African American (UM/CASE/NCAT) (U01AG052410, R01AG028786).

The four LSACs are: the Human Genome Sequencing Center at the Baylor College of Medicine (U54 HG003273), the Broad Institute Genome Center (U54HG003067), The American Genome Center at the Uniformed Services University of the Health Sciences (U01AG057659), and the

Washington University Genome Institute (U54HG003079).

Biological samples and associated phenotypic data used in primary data analyses were stored at Study Investigators institutions, and at the National Cell Repository for Alzheimer's Disease (NCRAD, U24AG021886) at Indiana University funded by NIA. Associated Phenotypic Data used in primary and secondary data analyses were provided by Study Investigators, the NIA funded Alzheimer's Disease Centers (ADCs), and the National Alzheimer's Coordinating Center (NACC, U01AG016976) and the National Institute on Aging Genetics of Alzheimer's Disease Data Storage Site (NIAGADS, U24AG041689) at the University of Pennsylvania, funded by NIA This research was supported in part by the Intramural Research Program of the National Institutes of health, National Library of Medicine. Contributors to the Genetic Analysis Data included Study Investigators on projects that were individually funded by NIA, and other NIH institutes, and by private U.S. organizations, or foreign governmental or nongovernmental organizations.

An up to date acknowledgment statement can be found on the ADSP site: https://www.niagads.org/adsp/content/acknowledgement-statement.

Data collection and sharing for this project was funded by the Alzheimer's Disease Neuroimaging Initiative (ADNI) (National Institutes of Health Grant U01 AG024904) and DOD ADNI (Department of Defense award number W81XWH-12-2-0012). ADNI is funded by the National Institute on Aging, the National Institute of Biomedical Imaging and Bioengineering, and through generous contributions from the following: AbbVie, Alzheimer's Association; Alzheimer's Drug Discovery Foundation; Araclon Biotech; BioClinica, Inc.; Biogen; Bristol-Myers Squibb Company; CereSpir, Inc.; Cogstate; Eisai Inc.; Elan Pharmaceuticals, Inc.; Eli Lilly and Company; EuroImmun; F. Hoffmann-La Roche Ltd and its affiliated company Genentech, Inc.; Fujirebio; GE Healthcare; IXICO Ltd.; Janssen Alzheimer Immunotherapy Research & Development, LLC.; Johnson & Johnson Pharmaceutical Research & Development LLC.; Lumosity; Lundbeck; Merck & Co., Inc.; Meso Scale Diagnostics, LLC.; NeuroRx Research; Neurotrack Technologies; Novartis Pharmaceuticals Corporation; Pfizer Inc.; Piramal Imaging; Servier; Takeda Pharmaceutical Company; and Transition Therapeutics. The Canadian Institutes of Health Research is providing funds to support ADNI clinical sites in Canada. Private sector contributions are facilitated by the Foundation for the National Institutes of Health (www.fnih.org). The grantee organization is the Northern California Institute for Research and Education, and the study is coordinated by the Alzheimer's Therapeutic Research Institute at the University of Southern California. ADNI data are disseminated by the Laboratory for Neuro Imaging at the University of Southern California.

Additional information to include in an acknowledgment statement can be found on the LONI site: https://adni.loni.usc.edu/wp-content/uploads/how_to_apply/ADNI_Data_Use_Agreement.pdf.

The Alzheimer's Disease Genetics Consortium (ADGC) supported sample preparation, whole exome sequencing and data processing through NIA grant U01AG032984. Sequencing data generation and harmonization is supported by the Genome Center for Alzheimer's Disease,

U54AG052427, and data sharing is supported by NIAGADS, U24AG041689. Samples from the National Centralized Repository for Alzheimer's Disease and Related Dementias (NCRAD), which receives government support under a cooperative agreement grant (U24 AG021886) awarded by the National Institute on Aging (NIA), were used in this study. We thank contributors who collected samples used in this study, as well as patients and their families, whose help and participation made this work possible. NIH grants supported enrollment and data collection for the individual studies including: GenerAAtions R01AG20688 (PI M. Daniele Fallin, PhD); Miami/Duke R01 AG027944, R01 AG028786 (PI Margaret A. Pericak-Vance, PhD); NC A&T P20 MD000546, R01 AG28786-01A1 (PI Goldie S. Byrd, PhD); Case Western (PI Jonathan L. Haines, PhD); MIRAGE R01 AG009029 (PI Lindsay A. Farrer, PhD); ROS P30AG10161, R01AG15819, R01AG30146, TGen (PI David A. Bennett, MD); MAP R01AG17917, R01AG15819, TGen (PI David A. Bennett, MD). The NACC database is funded by NIA/NIH Grant U01 AG016976. NACC data are contributed by the NIA-funded ADCs: P30 AG019610 (PI Eric Reiman, MD), P30 AG013846 (PI Neil Kowall, MD), P30 AG062428-01 (PI James Leverenz, MD) P50 AG008702 (PI Scott Small, MD), P50 AG025688 (PI Allan Levey, MD, PhD), P50 AG047266 (PI Todd Golde, MD, PhD), P30 AG010133 (PI Andrew Saykin, PsyD), P50 AG005146 (PI Marilyn Albert, PhD), P30 AG062421-01 (PI Bradley Hyman, MD, PhD), P30 AG062422-01 (PI Ronald Petersen, MD, PhD), P50 AG005138 (PI Mary Sano, PhD), P30 AG008051 (PI Thomas Wisniewski, MD), P30 AG013854 (PI Robert Vassar, PhD), P30 AG008017 (PI Jeffrey Kaye, MD), P30 AG010161 (PI David Bennett, MD), P50 AG047366 (PI Victor Henderson, MD, MS), P30 AG010129 (PI Charles DeCarli, MD), P50 AG016573 (PI Frank LaFerla, PhD), P30 AG062429-01(PI James Brewer, MD, PhD), P50 AG023501 (PI Bruce Miller, MD), P30 AG035982 (PI Russell Swerdlow, MD), P30 AG028383 (PI Linda Van Eldik, PhD), P30 AG053760 (PI Henry Paulson, MD, PhD), P30 AG010124 (PI John Trojanowski, MD, PhD), P50 AG005133 (PI Oscar Lopez, MD), P50 AG005142 (PI Helena Chui, MD), P30 AG012300 (PI Roger Rosenberg, MD), P30 AG049638 (PI Suzanne Craft, PhD), P50 AG005136 (PI Thomas Grabowski, MD), P30 AG062715-01 (PI Sanjay Asthana, MD, FRCP), P50 AG005681 (PI John Morris, MD), P50 AG047270 (PI Stephen Strittmatter, MD, PhD).

This work was supported by grants from the National Institutes of Health (R01AG044546, P01AG003991, RF1AG053303, R01AG058501, U01AG058922, RF1AG058501 and R01AG057777). The recruitment and clinical characterization of research participants at Washington University were supported by NIH P50 AG05681, P01 AG03991, and P01 AG026276. This work was supported by access to equipment made possible by the Hope Center for Neurological Disorders, and the Departments of Neurology and Psychiatry at Washington University School of Medicine.

We thank the contributors who collected samples used in this study, as well as patients and their families, whose help and participation made this work possible. Members of the National Institute on Aging Late-Onset Alzheimer Disease/National Cell Repository for Alzheimer Disease (NIA-LOAD NCRAD) Family Study Group include the following: Richard Mayeux, MD, MSc; Martin Farlow, MD; Tatiana Foroud, PhD; Kelley Faber, MS; Bradley F. Boeve, MD; Neill R. Graff-Radford, MD; David A. Bennett, MD; Robert A. Sweet, MD; Roger Rosenberg, MD; Thomas D. Bird, MD; Carlos Cruchaga, PhD; and Jeremy M. Silverman, PhD.

This work was partially supported by grant funding from NIH R01 AG039700 and NIH P50 AG005136. Subjects and samples used here were originally collected with grant funding from NIH U24 AG026395, U24 AG021886, P50 AG008702, P01 AG007232, R37 AG015473, P30 AG028377, P50 AG05128, P50 AG16574, P30 AG010133, P50 AG005681, P01 AG003991, U01MH046281, U01 MH046290 and U01 MH046373. The funders had no role in study design, analysis or preparation of the manuscript. The authors declare no competing interests.

This work was supported by the National Institutes of Health (R01 AG027944, R01 AG028786 to MAPV, R01 AG019085 to JLH, P20 MD000546); a joint grant from the Alzheimer's Association (SG-14-312644) and the Fidelity Biosciences Research Initiative to MAPV; the BrightFocus Foundation (A2011048 to MAPV). NIA-LOAD Family-Based Study supported the collection of samples used in this study through NIH grants U24 AG026395 and R01 AG041797 and the MIRAGE cohort was supported through the NIH grants R01 AG025259 and R01 AG048927. We thank contributors, including the Alzheimer's disease Centers who collected samples used in this study, as well as patients and their families, whose help and participation made this work possible. Study design: HNC, BWK, JLH, MAPV; Sample collection: MLC, JMV, RMC, LAF, JLH, MAPV; Whole exome sequencing and Sanger sequencing: SR, PLW; Sequencing data analysis: HNC, BWK, KLHN, SR, MAK, JRG, ERM, GWB, MAPV; Statistical analysis: BWK, KLHN, JMJ, MAPV; Preparation of manuscript: HNC, BWK. The authors jointly discussed the experimental results throughout the duration of the study. All authors read and approved the final manuscript.

Data collection and sharing for this project was supported by the Washington Heights-Inwood Columbia Aging Project (WHICAP, PO1AG07232, R01AG037212, RF1AG054023) funded by the National Institute on Aging (NIA) and by the National Center for Advancing Translational Sciences, National Institutes of Health, through Grant Number UL1TR001873. This manuscript has been reviewed by WHICAP investigators for scientific content and consistency of data interpretation with previous WHICAP Study publications. We acknowledge the WHICAP study participants and the WHICAP research and support staff for their contributions to this study.

This work was supported by grants from the National Institutes of Health (R01AG044546, P01AG003991, RF1AG053303, R01AG058501, U01AG058922, RF1AG058501 and R01AG057777). The recruitment and clinical characterization of research participants at Washington University were supported by NIH P50 AG05681, P01 AG03991, and P01 AG026276. This work was supported by access to equipment made possible by the Hope Center for Neurological Disorders, and the Departments of Neurology and Psychiatry at Washington University School of Medicine.

We thank the contributors who collected samples used in this study, as well as patients and their families, whose help and participation made this work possible. Members of the National Institute on Aging Late-Onset Alzheimer Disease/National Cell Repository for Alzheimer Disease (NIA-LOAD NCRAD) Family Study Group include the following: Richard Mayeux, MD, MSc; Martin Farlow, MD; Tatiana Foroud, PhD; Kelley Faber, MS; Bradley F. Boeve, MD;

Neill R. Graff-Radford, MD; David A. Bennett, MD; Robert A. Sweet, MD; Roger Rosenberg, MD; Thomas D. Bird, MD; Carlos Cruchaga, PhD; and Jeremy M. Silverman, PhD.

This work was supported by grants from the National Institutes of Health (R01AG044546, P01AG003991, RF1AG053303, R01AG058501, U01AG058922, RF1AG058501 and R01AG057777). The recruitment and clinical characterization of research participants at Washington University were supported by NIH P50 AG05681, P01 AG03991, and P01 AG026276. This work was supported by access to equipment made possible by the Hope Center for Neurological Disorders, and the Departments of Neurology and Psychiatry at Washington University School of Medicine.

We thank the contributors who collected samples used in this study, as well as patients and their families, whose help and participation made this work possible. Members of the National Institute on Aging Late-Onset Alzheimer Disease/National Cell Repository for Alzheimer Disease (NIA-LOAD NCRAD) Family Study Group include the following: Richard Mayeux, MD, MSc; Martin Farlow, MD; Tatiana Foroud, PhD; Kelley Faber, MS; Bradley F. Boeve, MD; Neill R. Graff-Radford, MD; David A. Bennett, MD; Robert A. Sweet, MD; Roger Rosenberg, MD; Thomas D. Bird, MD; Carlos Cruchaga, PhD; and Jeremy M. Silverman, PhD.

Mayo RNAseq Study- Study data were provided by the following sources: The Mayo Clinic Alzheimer's Disease Genetic Studies, led by Dr. Nilufer Ertekin-Taner and Dr. Steven G. Younkin, Mayo Clinic, Jacksonville, FL using samples from the Mayo Clinic Study of Aging, the Mayo Clinic Alzheimer's Disease Research Center, and the Mayo Clinic Brain Bank. Data collection was supported through funding by NIA grants P50 AG016574, R01 AG032990, U01 AG046139, R01 AG018023, U01 AG006576, U01 AG006786, R01 AG025711, R01 AG017216, R01 AG003949, NINDS grant R01 NS080820, CurePSP Foundation, and support from Mayo Foundation. Study data includes samples collected through the Sun Health Research Institute Brain and Body Donation Program of Sun City, Arizona. The Brain and Body Donation Program is supported by the National Institute of Neurological Disorders and Stroke (U24 NS072026 National Brain and Tissue Resource for Parkinson's Disease and Related Disorders), the National Institute on Aging (P30 AG19610 Arizona Alzheimer's Disease Core Center), the Arizona Department of Health Services (contract 211002, Arizona Alzheimer's Research Center), the Arizona Biomedical Research Commission (contracts 4001, 0011, 05-901 and 1001 to the Arizona Parkinson's Disease Consortium) and the Michael J. Fox Foundation for Parkinson's Research

ROSMAP- We are grateful to the participants in the Religious Order Study, the Memory and Aging Project. This work is supported by the US National Institutes of Health [U01 AG046152, R01 AG043617, R01 AG042210, R01 AG036042, R01 AG036836, R01 AG032990, R01 AG18023, RC2 AG036547, P50 AG016574, U01 ES017155, KL2 RR024151, K25 AG041906-01, R01 AG30146, P30 AG10161, R01 AG17917, R01 AG15819, K08 AG034290, P30 AG10161 and R01 AG11101.

Mount Sinai Brain Bank (MSBB)- This work was supported by the grants R01AG046170, RF1AG054014, RF1AG057440 and R01AG057907 from the NIH/National Institute on Aging (NIA). R01AG046170 is a component of the AMP-AD Target Discovery and Preclinical

Validation Project. Brain tissue collection and characterization was supported by NIH HHSN271201300031C.

This study was supported by the National Institute on Aging (NIA) grants AG030653, AG041718, AG064877 and P30-AG066468.

We would like to thank study participants, their families, and the sample collectors for their invaluable contributions. This research was supported in part by the National Institute on Aging grant U01AG049508 (PI Alison M. Goate). This research was supported in part by Genentech, Inc. (PI Alison M. Goate, Robert R. Graham).

The NACC database is funded by NIA/NIH Grant U01 AG016976. NACC data are contributed by these NIA-funded ADCs: P30 AG013846 (PI Neil Kowall, MD), P50 AG008702 (PI Scott Small, MD), P50 AG025688 (PI Allan Levey, MD, PhD), P30 AG010133 (PI Andrew Saykin, PsyD), P50 AG005146 (PI Marilyn Albert, PhD), P50 AG005134 (PI Bradley Hyman, MD, PhD), P50 AG016574 (PI Ronald Petersen, MD, PhD), P30 AG013854 (PI M. Marsel Mesulam, MD), P30 AG008017 (PI Jeffrey Kaye, MD), P30 AG010161 (PI David Bennett, MD), P30 AG010129 (PI Charles DeCarli, MD), P50 AG016573 (PI Frank LaFerla, PhD), P50 AG005131 (PI Douglas Galasko, MD), P30 AG028383 (PI Linda Van Eldik, PhD), P30 AG010124 (PI John Trojanowski, MD, PhD), P50 AG005142 (PI Helena Chui, MD), P30 AG012300 (PI Roger Rosenberg, MD), P50 AG005136 (PI Thomas Grabowski, MD), P50 AG005681 (PI John Morris, MD), P30 AG028377 (Kathleen Welsh-Bohmer, PhD), and P50 AG008671 (PI Henry Paulson, MD, PhD).

Samples from the National Cell Repository for Alzheimer's Disease (NCRAD), which receives government support under a cooperative agreement grant (U24 AG21886) awarded by the National Institute on Aging (NIA), were used in this study. We thank contributors who collected samples used in this study, as well as patients and their families, whose help and participation made this work possible.

The Alzheimer's Disease Genetics Consortium supported the collection of samples used in this study through National Institute on Aging (NIA) grants U01AG032984 and RC2AG036528.

We acknowledge the generous contributions of the Cache County Memory Study participants. Sequencing for this study was funded by RF1AG054052 (PI: John S.K. Kauwe)

**Acknowledgments for the use of GWAS data distributed by NIAGADS**

The NIA Genetics of Alzheimer's Disease Data Storage Site (NIAGADS) is supported by a collaborative agreement from the National Institute on Aging, U24AG041689.

NG00047: The NIA supported this work through grants U01-AG032984, RC2-AG036528, U01-AG016976 (Dr Kukull); U24 AG026395, U24 AG026390, R01AG037212, R37 AG015473 (Dr Mayeux); K23AG034550 (Dr Reitz); U24-AG021886 (Dr Foroud); R01AG009956, RC2 AG036650 (Dr Hall); UO1 AG06781, UO1 HG004610 (Dr Larson); R01 AG009029 (Dr Farrer); 5R01AG20688

(Dr Fallin); P50 AG005133, AG030653 (Dr Kamboh); R01 AG019085 (Dr Haines); R01 AG1101, R01 AG030146, RC2 AG036650 (Dr Evans); P30AG10161, R01AG15819, R01AG30146, R01AG17917, R01AG15819 (Dr Bennett); R01AG028786 (Dr Manly); R01AG22018, P30AG10161 (Dr Barnes); P50AG16574 (Dr Ertekin-Taner, Dr Graff-Radford), R01 AG032990 (Dr Ertekin-Taner), KL2 RR024151 (Dr Ertekin-Taner); R01 AG027944, R01 AG028786 (Dr Pericak-Vance); P20 MD000546, R01 AG28786-01A1 (Dr Byrd); AG005138 (Dr Buxbaum); P50 AG05681, P01 AG03991, P01 AG026276 (Dr Goate); and P30AG019610, P30AG13846, U01-AG10483, R01CA129769, R01MH080295, R01AG017173, R01AG025259, R01AG33193, P50AG008702, P30AG028377, AG05128, AG025688, P30AG10133, P50AG005146, P50AG005134, P01AG002219, P30AG08051, MO1RR00096, UL1RR029893, P30AG013854, P30AG008017, R01AG026916, R01AG019085, P50AG016582, UL1RR02777, R01AG031581, P30AG010129, P50AG016573, P50AG016575, P50AG016576, P50AG016577, P50AG016570, P50AG005131, P50AG023501, P50AG019724, P30AG028383, P50AG008671, P30AG010124, P50AG005142, P30AG012300, AG010491, AG027944, AG021547, AG019757, P50AG005136 (Alzheimer Disease GeneticsConsortium [ADGC]). We thank Creighton Phelps, Stephen Synder, and Marilyn Miller from the NIA, who are ex-officio members of the ADGC. Support was also provided by the Alzheimer's Association (IIRG-08-89720 [Dr Farrer] and IIRG-05-14147 [Dr Pericak-Vance]), National Institute of Neurological Disorders and Stroke grant NS39764, National Institute of Mental Health grant MH60451, GlaxoSmithKline, and the Office of Research and Development, Biomedical Laboratory Research Program, US Department of Veterans Affairs Administration. For the ADGC, biological samples and associated phenotypic data used in primary data analyses were stored at principal investigators' institutions and at the National Cell Repository for Alzheimer's Disease (NCRAD) at Indiana University, funded by the NIA. Associated phenotypic data used in secondary data analyses were stored at the National Alzheimer's Coordinating Center and at the NIA Alzheimer's Disease Data Storage Site at the University of Pennsylvania, funded by the NIA. Contributors to the genetic analysis data included principal investigators on projects ndividually funded by the NIA, other NIH institutes, or private entities.

**Acknowledgments for the use of MARS and LATC**

We thank all Minority Aging Research Study and Latino Core participants and the Rush Alzheimer's Disease Center staff. This database was funded by the NIH/NIA grants R01AG22018 (MARS) and P30AG 072975 (ADC).

**Acknowledgments for other GWAS and phenotype data**

The NACC database is funded by NIA/NIH Grant U01 AG016976. NACC data are contributed by the NIA-funded ADCs: P30 AG019610 (PI Eric Reiman, MD), P30 AG013846 (PI Neil Kowall, MD), P30 AG062428-01 (PI James Leverenz, MD) P50 AG008702 (PI Scott Small, MD), P50 AG025688 (PI Allan Levey, MD, PhD), P50 AG047266 (PI Todd Golde, MD, PhD), P30 AG010133 (PI Andrew Saykin, PsyD), P50 AG005146 (PI Marilyn Albert, PhD), P30 AG062421-01 (PI Bradley Hyman, MD, PhD), P30 AG062422-01 (PI Ronald Petersen, MD, PhD), P50 AG005138 (PI Mary Sano, PhD), P30 AG008051 (PI Thomas Wisniewski, MD), P30 AG013854 (PI Robert Vassar, PhD), P30 AG008017 (PI Jeffrey Kaye, MD), P30 AG010161 (PI David Bennett, MD), P50 AG047366 (PI Victor Henderson, MD, MS), P30 AG010129 (PI Charles DeCarli, MD), P50 AG016573 (PI Frank

LaFerla, PhD), P30 AG062429-01(PI James Brewer, MD, PhD), P50 AG023501 (PI Bruce Miller, MD), P30 AG035982 (PI Russell Swerdlow, MD), P30 AG028383 (PI Linda Van Eldik, PhD), P30 AG053760 (PI Henry Paulson, MD, PhD), P30 AG010124 (PI John Trojanowski, MD, PhD), P50 AG005133 (PI Oscar Lopez, MD), P50 AG005142 (PI Helena Chui, MD), P30 AG012300 (PI Roger Rosenberg, MD), P30 AG049638 (PI Suzanne Craft, PhD), P50 AG005136 (PI Thomas Grabowski, MD), P30 AG062715-01 (PI Sanjay Asthana, MD, FRCP), P50 AG005681 (PI John Morris, MD), P50 AG047270 (PI Stephen Strittmatter, MD, PhD).

The genotypic and associated phenotypic data used in the study "Multi-Site Collaborative Study for Genotype-Phenotype Associations in Alzheimer's Disease (GenADA)" were provided by the GlaxoSmithKline, R&D Limited.

ROSMAP study data were provided by the Rush Alzheimer's Disease Center, Rush University Medical Center, Chicago. Data collection was supported through funding by NIA grants P30AG10161, R01AG15819, R01AG17917, R01AG30146, R01AG36836, U01AG32984, U01AG46152, the Illinois Department of Public Health, and the Translational Genomics Research Institute.

The AddNeuroMed data are from a public-private partnership supported by EFPIA companies and SMEs as part of InnoMed (Innovative Medicines in Europe), an Integrated Project funded by the European Union of the Sixth Framework program priority FP6-2004-LIFESCIHEALTH-5. Clinical leads responsible for data collection are Iwona Kłoszewska (Lodz), Simon Lovestone (London), Patrizia Mecocci (Perugia), Hilkka Soininen (Kuopio), Magda Tsolaki (Thessaloniki), and Bruno Vellas (Toulouse), imaging leads are Andy Simmons (London), Lars-Olad Wahlund (Stockholm) and Christian Spenger (Zurich) and bioinformatics leads are Richard Dobson (London) and Stephen Newhouse (London).

### Supplementary Note

##### *APOE* GENOTYPE ASCERTAINMENT

We directed specific attention to the genotyping of the SNPs determining the main *APOE* genotype (rs429358 and rs7412), rs769455-T (*APOE*[R145C]), and rs376170967-A (*APOE*[R150H]). Note that Arg145Cys (R145C) is also sometimes referred to as Arg163Cys when the first 18 codons of *APOE* encoding a signal peptide are included; and respectively Arg150Cys is also referred to as Arg168Cys. The rs429358 and rs7412 alternate (minor) alleles were respectively used to determine the number of *APOE ε4* and *APOE ε2* alleles. The *ε3* allele was determined by a reference (major) allele call at both rs429359 and rs7412. Determining the *ε2*/*ε4* genotype would in theory require phase information (i.e., knowing whether alleles are observed on the same chromosome copy) to distinguish it from *ε1*/*ε3*, but since the *ε1* allele is extremely rare[1], we followed common practice to assign the *ε2*/*ε4* genotype. We considered the two triplets of variants separately (rs429358, rs7412, rs769455-T) and (rs429358, rs7412, rs376170967-A). Hereafter, we described the rs769455-T triplet genotyping and the same protocol was applied for the rs376170967-A triplet genotyping.

<u>In the discovery dataset</u>, composed of WGS and WES data, the three variants were directly called in the sequencing data distributed by NIAGADS (**Table S13).** In ADSP WGS, the three variants showed low genotype missingness across subjects (2-3%). We therefore decided to simply exclude subjects in ADSP WGS with missing genotypes for any of the three variants. In ADSP WES, there was a high genotype missingness at rs7412 (32.5%). This resulted from a low read depth and genotype quality in some of the different WES capture kits that were used in the ADSP WES project[2]. We therefore sought to re-call all three variants in order to fill out missing information where possible. We inferred the variants' genotype if one of the following two conditions was met: (i) read depth (DP) and genotype quality (GQ) were, respectively, greater than or equal to 6 and 20, observing at least 20% alternate allele reads to call a heterozygote (e.g. *ε2*/*ε3*); or (ii) considering only DP greater than or equal to 4 with, either, all calls corresponding to one allele to call a homozygote (e.g. *ε4*/*ε4*), or, each allele called on at least 40% of the reads, to call a heterozygote. Importantly, as a quality check, using these thresholds, we did not observe any discordance in the inferred *APOE* genotype across 3499 duplicates between

WGS and WES. After this first round of *APOE* genotype ascertainment, some individuals still had either the rs7412 or rs429358 genotype missing (i.e., only one of the two variants could be called using the above criteria), making it impossible to infer their *APOE* genotype from WES data alone. 110 individuals (3.4%) included in the discovery analysis were in this situation (**Figure S3**). In order to determine the main *APOE* genotype In these 110 individuals, we used the following approach. Many of these individuals had a reported *APOE* genotype in their demographics that could be used to complete the missing information in a second additional round of *APOE* genotype ascertainment. This approach was preferred over relying solely on the *APOE* genotype in the demographics, since the genotype calls on the WES data are expected to provide higher accuracy compared to other commonly used *APOE* genotyping methods[3]. Here, it is worth noting that the standard read length of the WES data (100 to 150 bp) entails that rs769455 will often be on the same read as rs7412 (39 bp apart) and in rarer instances on the same read as rs429358 (99 bp apart). Thus, for this second ascertainment round, to avoid observing only one of the chromosome 19 pair in the sequencing reads at rs769455, we required rs769455 and other available APOE variants to meet criteria (i) from above. To illustrate, consider the example where an individual in the sequencing data was homozygous for the reference allele at rs429358 but had a missing genotype at rs7412. In this case, from the WES information, we know that this individual is not carrying an *ε4* allele, but we cannot determine the presence or absence of an *ε2* allele. We then turned to the information from the *APOE* genotype reported in the demographics to infer the most likely *APOE* genotype. In our particular example, if the individual has a reported *APOE* genotype *ε3/ε3, ε2/ε3,* or *ε2/ε2* then the information in the WES data is deemed concordant with the reported *APOE* and we used the reported *APOE*. However, if the reported *APOE* genotype was *ε4/ε4, ε3/ε4* then we would correct this genotype to *ε3/ε3*, based on the WES information that clearly indicated there were no APOE*ε4* reads. This can be generalized as: (a) for individuals with high DP at rs429358 and for whom resolving the *APOE* genotype simply required changing *ε3* to *ε4* or vice-versa, then the information from the WES at rs429358 was used, (b) similarly for individuals with high DP at rs7412 and for whom resolving *APOE* genotype simply required changing *ε3* to *ε2* or vice-versa the information from the WES at rs7412 was used. Finally, subjects who had no call at rs7412 and rs429358 in the sequencing data were excluded. **Figure S3** provides a graphical representation of this workflow and provides the exact number of individuals in each category. Additionally, sensitivity analyses, in which we restricted the analysis to ADSP WES individuals who had direct calls at all 3 APOE missense variants, emphasize that the significance of our results and effect sizes remain unchanged (**Table S14**).

In the replication dataset, which did not contain individuals with next generation sequencing data, the main *APOE* genotype was prioritized in the following order: (i) directly obtained from the microarray genotyping, (ii) provided with the primary study demographics (genotyping methods described elsewhere[4,5]), (iii) imputed with high confidence ($r^2 > 0.8$). rs769455_T was directly genotyped on the Exome Chip microarrays and was imputed in other datasets (cf Imputation section in the main text).

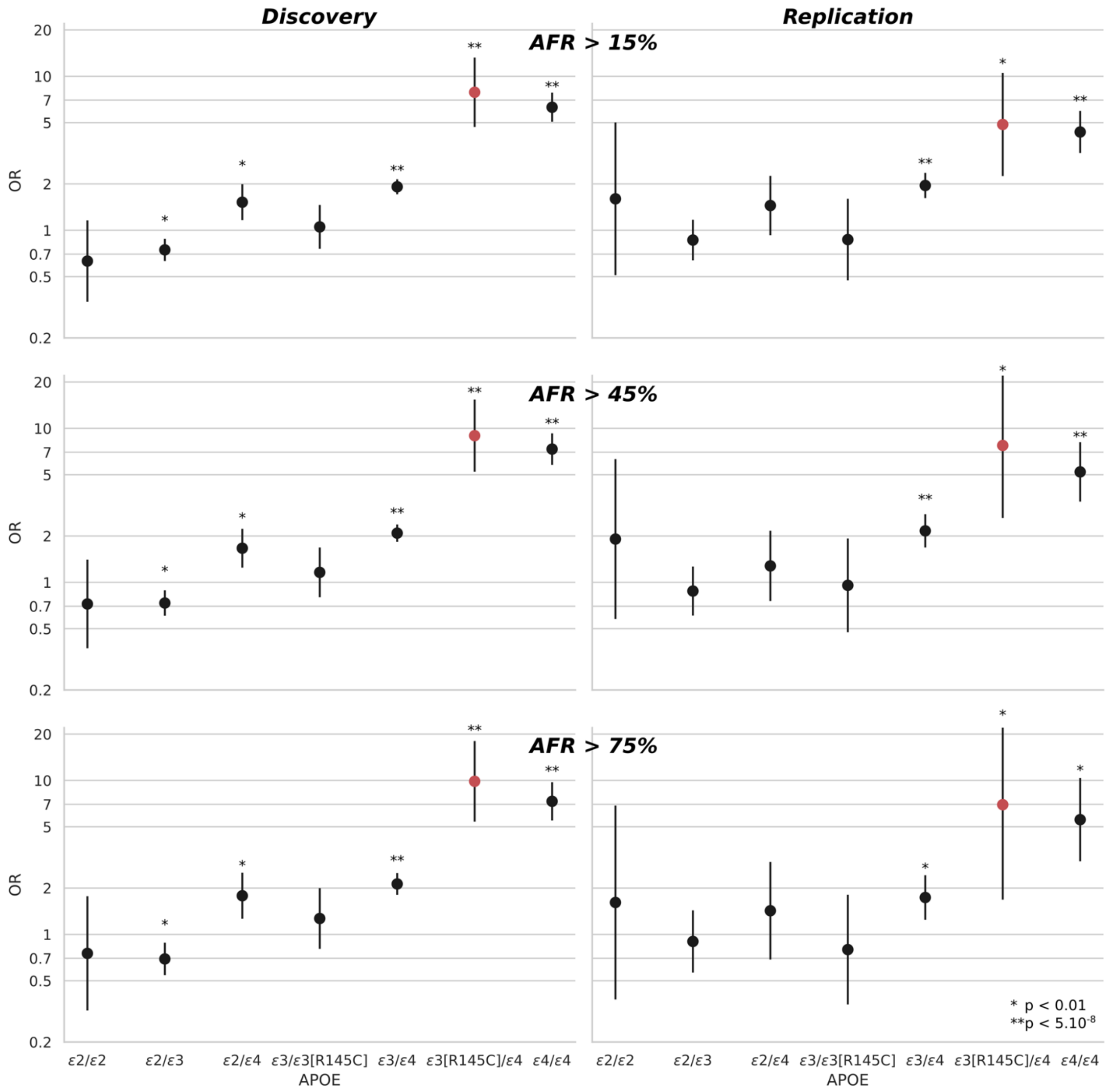


**Figure S1. *APOEε3*[R145C]/*ε4* individuals have an AD risk comparable to *APOEε4/ε4* individuals regardless of the African ancestry cutoff.** Sensitivity analysis for the AD risk odds ratio (OR) per *APOE* group in the discovery and replication for several African ancestry cutoffs (15%, 45%, and 75%). AD risk per *APOE* group assessed compared to the *APOEε3/ε3* reference group (i.e., $OR_{APOE\varepsilon3/\varepsilon3} = 1$) in our discovery sample (left column) composed of next generation sequencing data from the ADSP dataset, and in our replication (right column) composed of microarray data imputed on the TOPMed reference panel.

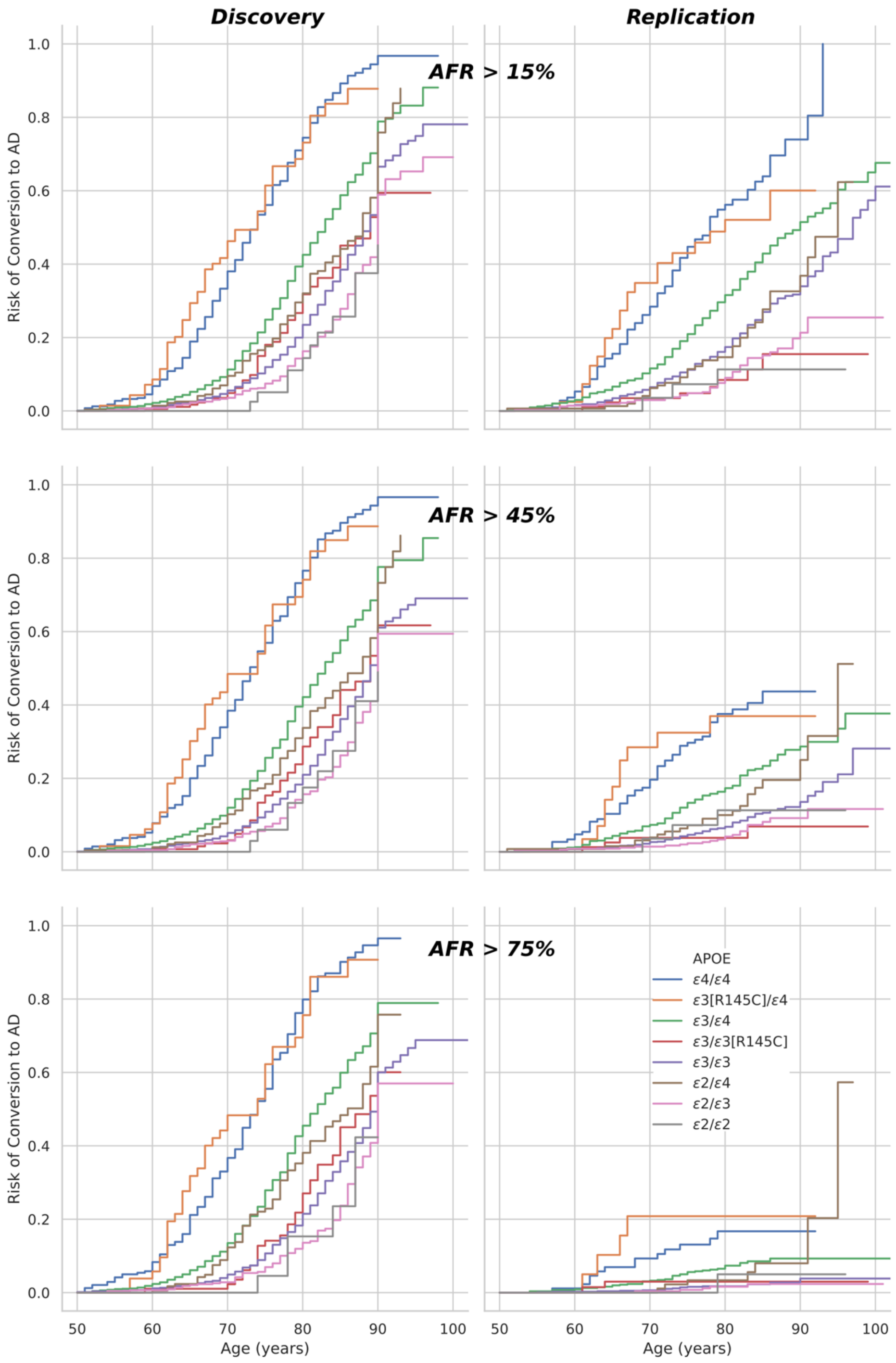


**Figure S2. The *APOEε3/*[R145C]/*ε4* cumulative incidence closely mirrors the *APOEε4/ε4* cumulative incidence regardless of the African ancestry cutoff.** Sensitivity analysis for competing risk regression per *APOE* group in the discovery and replication for several African ancestry cutoffs (15%, 45%, and 75%).

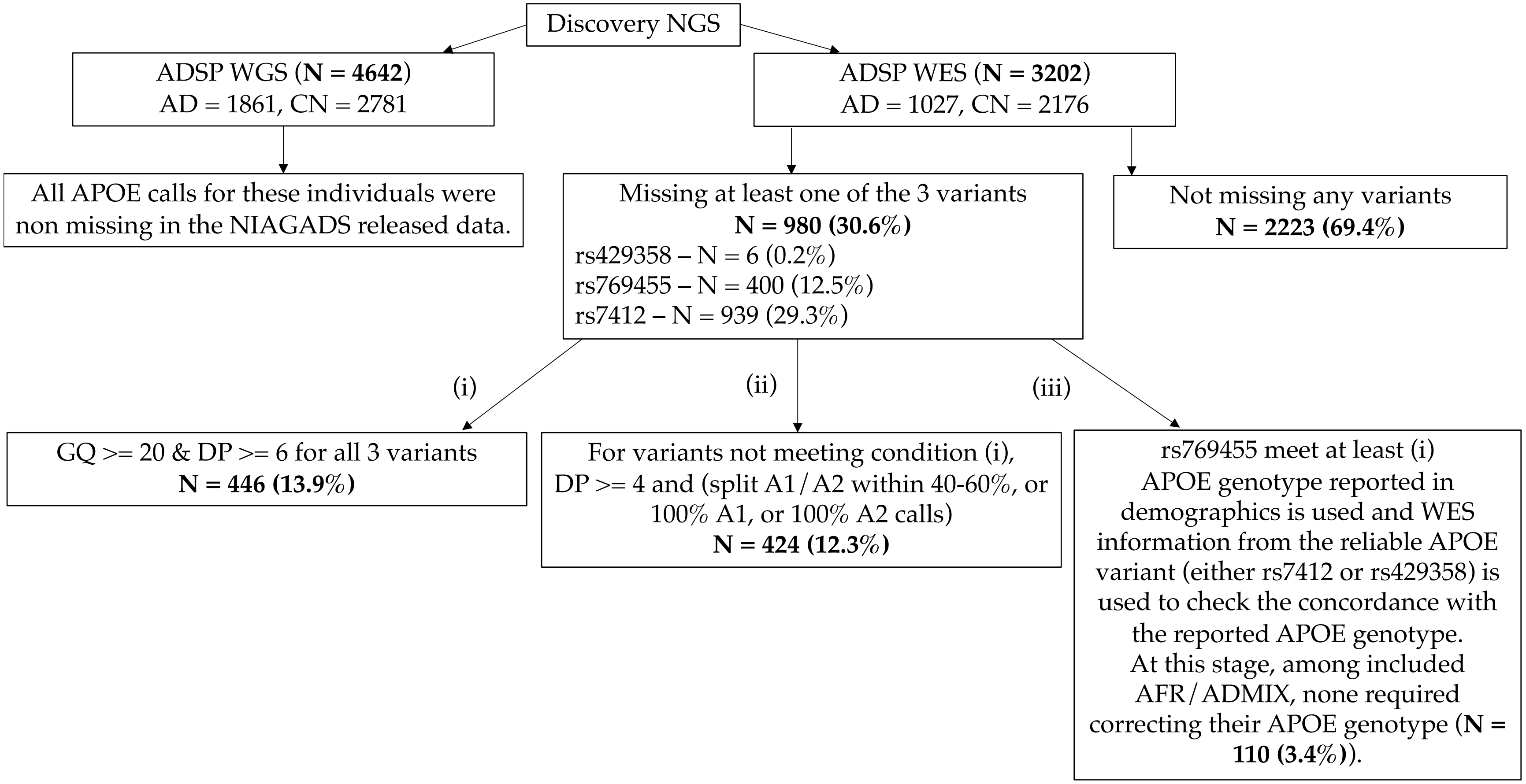


**Figure S3. Flowchart describing the *APOE* alleles and R145C genotyping among African and Admixed-African individuals included in the discovery analysis.**

**Table S1. Missense variants on *APOE* canonical transcript reported in gnomADv.3.1**. Pos: position on chromosome 19 in build hg38, AF: alternate allele frequency, AC: allele count, AN: allele number, Hom: Homozygote count. Only the first ten missense variants in term of allele frequency in gnomAD are reported here (AC > 20).

| | | | | HGVs | | Overall gnomAD.v3.1 | | | | African/African-American | | | | Latino/Admixed American | | | | European (non-Finnish) | | | |
|---|---|---|---|---|---|---|---|---|---|---|---|---|---|---|---|---|---|---|---|---|---|
| **Pos (hg38)** | **rsIDs** | **Ref** | **Alt** | **new** | **standard** | **AC** | **AN** | **AF** | **Hom** | **AC** | **AN** | **AF** | **Hom** | **AC** | **AN** | **AF** | **Hom** | **AC** | **AN** | **AF** | **Hom** |
| 44908684 | rs429358 | T | C | p.Cys130Arg | p.Cys112Arg | 23875 | 151972 | 0.157101 | 1998 | 8917 | 41390 | 0.215439 | 950 | 1718 | 15280 | 0.112435 | 90 | 9371 | 67924 | 0.137963 | 676 |
| 44908822 | rs7412 | C | T | p.Arg176Cys | p.Arg158Cys | 11838 | 152004 | 0.07788 | 546 | 4335 | 41402 | 0.104705 | 247 | 640 | 15262 | 0.041934 | 17 | 5401 | 67956 | 0.079478 | 224 |
| 44908783 | rs769455 | C | T | p.Arg163Cys | p.Arg145Cys | 978 | 152126 | 0.006429 | 15 | 869 | 41444 | 0.020968 | 15 | 83 | 15274 | 0.005434 | 0 | 5 | 67996 | 7.35E-05 | 0 |
| 44907853 | rs769452 | T | C | p.Leu46Pro | p.Leu28Pro | 293 | 152188 | 0.001925 | 0 | 11 | 41454 | 0.000265 | 0 | 14 | 15282 | 0.000916 | 0 | 168 | 68034 | 0.002469 | 0 |
| 44909057 | rs199768005 | T | A | p.Val254Glu | p.Val236Glu | 72 | 152080 | 0.000473 | 0 | 16 | 41424 | 0.000386 | 0 | 0 | 15272 | 0 | 0 | 52 | 67984 | 0.000765 | 0 |
| 44907807 | rs201672011 | G | A | p.Glu31Lys | p.Glu13Lys | 72 | 152186 | 0.000473 | 0 | 10 | 41458 | 0.000241 | 0 | 48 | 15282 | 0.003141 | 0 | 5 | 68024 | 7.35E-05 | 0 |
| 44908799 | rs376170967 | G | A | p.Arg168His | p.Arg150His | 62 | 152122 | 0.000408 | 0 | 58 | 41452 | 0.001399 | 0 | 3 | 15262 | 0.000197 | 0 | 1 | 67994 | 1.47E-05 | 0 |
| 44909101 | rs267606661 | C | G | p.Arg269Gly | p.Arg251Gly | 46 | 152200 | 0.000302 | 0 | 5 | 41450 | 0.000121 | 0 | 7 | 15280 | 0.000458 | 0 | 33 | 68022 | 0.000485 | 0 |
| 44908730 | rs267606664 | G | A | p.Gly145Asp | p.Gly127Asp | 22 | 152152 | 0.000145 | 0 | 3 | 41458 | 7.24E-05 | 0 | 1 | 15284 | 6.54E-05 | 0 | 17 | 67992 | 0.00025 | 0 |
| 44908915 | rs749750245 | C | T | p.Arg207Cys | p.Arg189Cys | 21 | 151884 | 0.000138 | 1 | 0 | 41406 | 0 | 0 | 21 | 15254 | 0.001377 | 1 | 0 | 67920 | 0 | 0 |

**Table S2. Queried cohort overview to identify Admixed and African ancestry individuals.**

| Cohort/Project | Genotyping Platform | Cohort-Platform ID | Sample (N) | Data Repository and Access ID |
|---|---|---|---|---|
| ADSP WES | Whole Exome Sequencing | ADSP_WES | 20503 | NIAGADS DSS (NG00067.v3) / NACC |
| ADSP WGS | Whole Genome Sequencing | ADSP_WGS | 16906 | NIAGADS DSS (NG00067.v5) / NACC |
| ACT | Illumina Human 660W-Quad | ACT | 2790 | NIAGADS (NG00034) / dbGaP (phs000234) |
| ADC1 | Illumina Human 660W-Quad | ADC1 | 2731 | NIAGADS (NG00022) / NACC |
| ADC2 | Illumina Human 660W-Quad | ADC2 | 928 | NIAGADS (NG00023) / NACC |
| ADC3 | Illumina Human OmniExpress | ADC3 | 1526 | NIAGADS (NG00024) / NACC |
| ADC4 | Illumina Human OmniExpress | ADC4 | 1054 | NIAGADS (NG00068) / NACC |
| ADC5 | Illumina Human OmniExpress | ADC5 | 1224 | NIAGADS (NG00069) / NACC |
| ADC6 | Illumina Human OmniExpress | ADC6 | 1333 | NIAGADS (NG00070) / NACC |
| ADC7 | Illumina Infinium Human OmniExpressExome | ADC7 | 1462 | NIAGADS (NG00071) / NACC |
| ADDNEUROMED | Illumina Human 610-Quad | ADM_Q | 315 | Synapse AddNeuroMed (syn4907804) |
| | Illumina Human OmniExpress | ADM_O | 329 | Synapse AddNeuroMed (syn4907804) |
| ADGC-ExomeChip | Illumina HumanExome BeadChip v1.0 at CHOP | CHOP | 5180 | NIAGADS (NG00081) / NACC |
| | Illumina HumanExome BeadChip v1.0 at Miami | MIA | 1923 | NIAGADS (NG00080) / NACC |
| | Illumina HumanExome BeadChip v1.0 at Northshore | NS | 5998 | NIAGADS (NG00079) / NACC |
| | Illumina HumanExome BeadChip v1.0 at WashU | WU | 868 | NIAGADS (NG00085) / NACC |
| ADNI | Illumina Human 610-Quad | ADNI_Q | 757 | LONI ADNI |
| | Illumina Human OmniExpress | ADNI_OE | 361 | LONI ADNI |
| | Illumina Omni 2.5 | ADNI_O25 | 812 | LONI ADNI |
| | Illumina Human OmniExpress | ADNI_DOD | 204 | LONI ADNIDOD |
| ADNI3 | Illumina Global Screening Array (GSA) | ADNI3 | 327 | LONI ADNI |
| IIDP African Americans | Illumina Human 1M-Duo | IIDP_AA | 1175 | NIAGADS (NG00047) |
| IIDP Yorubans | Illumina Human 1M-Duo | IIDP_YOR | 1264 | NIAGADS (NG00047) / cf. gaaindata.org/partner/IIDP |
| CIDR | Illumina Human Omni1-Quad | CIDR | 3101 | NIAGADS (NG00015) / dbGAP (phs000160) |
| GenADA | Affymetrix 500K | GSK | 1571 | dbGaP (phs000219) |
| LATC | Illumina Multi-Ethnic – BU | LATC | 63 | RADC Rush / Latino CORE Study |
| NIA-LOAD | Illumina Human 610-Quad | LOAD | 5220 | NIAGADS (NG00020) |

| | | | | |
|---|---|---|---|---|
| MARS | Illumina Multi-Ethnic – BU | MARS | 708 | RADC Rush / Minority Aging Research Study |
| MAYO | Illumina Human Hap300 | MAYO_1 | 2099 | Synapse AMP-AD (syn5591675) |
| MAYO2 | Illumina Omni 2.5 | MAYO_2 | 314 | Synapse AMP-AD (syn5550404) |
| MIRAGE | Illumina Human CNV370-Duo | MIRAGE_370 | 397 | NIAGADS (NG00031) |
| | Illumina Human 610-Quad | MIRAGE_610 | 1105 | NIAGADS (NG00031) |
| MTC | Illumina Human OmniExpress | MTC | 542 | NIAGADS (NG00096) |
| OHSU | Illumina Human CNV370-Duo | OHSU | 647 | NIAGADS (NG00017) |
| ROSMAP | Affymetrix GeneChip 6.0 - Broad Institute | ROSMAP_1B | 1126 | RADC Rush / Synapse AMP-AD (syn3219045) |
| | Affymetrix GeneChip 6.0 - TGen | ROSMAP_1T | 582 | RADC Rush / Synapse AMP-AD (syn3219045) |
| | Illumina Human OmniExpress 12 - Chop | ROSMAP_2C | 382 | RADC Rush / Synapse AMP-AD (syn7824841) |
| | Illumina Multi-Ethnic - BU | ROSMAP_3BU | 494 | RADC Rush |
| TARCC | Affymetrix 6.0 | TARCC | 2718 | NIAGADS (NG00097) / TARCC study |
| TGEN2 | Affymetrix 6.0 | TGEN | 1599 | NIAGADS (NG00028) |
| UPITT | Illumina Human Omni1-Quad | UPITT | 2440 | NIAGADS (NG00026) |
| UM-VU-MSSM | Illumina Human 1M-Duo, Illumina 1M | UVM_A | 1153 | NIAGADS (NG00042) |
| | Affymetrix 6.0 | UVM_B | 864 | NIAGADS (NG00042) |
| | Illumina Human 550K. Illumina Human 610-Quad | UVM_C | 445 | NIAGADS (NG00042) |
| WASHU | Illumina Human 610-Quad | WASHU_1 | 670 | NIAGADS (NG00030) |
| WASHU2 | Illumina Human OmniExpress | WASHU_2 | 235 | NIAGADS (NG00087) |
| WHICAP | Illumina Human OmniExpress | WHICAP | 647 | NIAGADS (NG00093) |

**Table S3. Overview of Alzheimer's disease sequencing project (ADSP) studies with whole-exome sequencing (WES) and/or whole-genome sequencing (WGS) available at NIAGADS DSS (NG00067).**

| Study | Accession Number | Related Datasets |
|---|---|---|
| Accelerating Medicines Partnership- Alzheimer's Disease (AMP-AD) | sa000011 | NG00067 – ADSP Umbrella |
| Cache County Study | sa000014 | NG00067 – ADSP Umbrella |
| University of Pittsburgh- Kamboh WGS | sa000012 | NG00067 – ADSP Umbrella |
| CurePSP and Tau Consortium PSP WGS | sa000016 | NG00067 – ADSP Umbrella |
| NIH, CurePSP and Tau Consortium PSP WGS | sa000015 | NG00067 – ADSP Umbrella |
| UCLA Progressive Supranuclear Palsy | sa000017 | NG00067 – ADSP Umbrella |
| NACC Genentech WGS | sa000013 | NG00067 – ADSP Umbrella |
| Alzheimer's Disease Sequencing Project (ADSP) | sa000001 | NG00067 – ADSP Umbrella |
| Alzheimer's Disease Neuroimaging Initiative (ADNI) | sa000002 | NG00067 – ADSP Umbrella |
| Alzheimer's Disease Genetics Consortium: African Americans (ADGC AA) | sa000003 | NG00067 – ADSP Umbrella |
| The Familial Alzheimer Sequencing (FASe) project | sa000004 | NG00067 – ADSP Umbrella |
| Brkanac – Family-based genome scan for AAO of LOAD | sa000005 | NG00067 – ADSP Umbrella |
| HIHG Miami Families with AD | sa000006 | NG00067 – ADSP Umbrella |
| Washington Heights/Inwood Columbia Aging Project (WHICAP) | sa000007 | NG00067 – ADSP Umbrella |
| Charles F. and Joanne Knight Alzheimer's Disease Research Center (Knight ADRC) | sa000008 | NG00067 – ADSP Umbrella |
| Corticobasal degeneration Study (CBD) | sa000009 | NG00067 – ADSP Umbrella |
| Progressive Supranuclear Palsy Study (PSP) | sa000010 | NG00067 – ADSP Umbrella |

**Table S4. Demographics of all the queried cohorts.** AFR: African, AMR: American (central and south; admixed), EAS: East Asian, SAS South Asian, EUR: European, otherwise ADMIX: admixed of these super ancestry categories.

| | Cohort | N total | Ancestry | | | | | | Diagnosis | | Sex - Females | | Age | |
|---|---|---|---|---|---|---|---|---|---|---|---|---|---|---|
| | | | AFR N | ADMIX N | AMR N | EAS N | SAS N | EUR N | CN N | AD N | CN N(%) | AD N(%) | CN μ(σ) | AD μ(σ) |
| *Discovery* | ADSP WES | 20503 | 3171 | 3174 | 125 | 7 | 2 | 14024 | 9617 | 8723 | 6101(63.4) | 5394(61.8) | 82.0(8.5) | 75.7(8.8) |
| | ADSP WGS | 16906 | 2240 | 4012 | 58 | 68 | 19 | 10509 | 6717 | 6434 | 4510(67.1) | 3896(60.6) | 78.2(8.5) | 74.1(10.5) |
| *Replication* | ACT | 2790 | 70 | 64 | 7 | 73 | 0 | 2576 | 1833 | 713 | 1000(54.6) | 462(64.8) | 82.9(6.5) | 82.1(6.6) |
| | ADC1 | 2731 | 92 | 58 | 47 | 20 | 0 | 2514 | 603 | 1946 | 354(58.7) | 1039(53.4) | 79.8(10.8) | 70.7(9.5) |
| | ADC2 | 928 | 0 | 2 | 0 | 0 | 0 | 926 | 124 | 707 | 87(70.2) | 366(51.8) | 80.1(9.2) | 72.9(7.1) |
| | ADC3 | 1526 | 0 | 5 | 0 | 0 | 0 | 1521 | 482 | 858 | 305(63.3) | 468(54.5) | 79.6(9.6) | 72.5(10.3) |
| | ADC4 | 1054 | 6 | 10 | 1 | 0 | 0 | 1037 | 420 | 452 | 257(61.2) | 237(52.4) | 79.2(8.7) | 72.6(9.0) |
| | ADC5 | 1224 | 0 | 1 | 0 | 0 | 0 | 1223 | 579 | 415 | 376(64.9) | 226(54.5) | 82.0(8.9) | 74.1(8.7) |
| | ADC6 | 1333 | 0 | 2 | 0 | 0 | 0 | 1331 | 352 | 567 | 238(67.6) | 304(53.6) | 80.1(8.9) | 66.9(12.0) |
| | ADC7 | 1462 | 0 | 4 | 0 | 0 | 0 | 1458 | 763 | 536 | 493(64.6) | 281(52.4) | 78.0(7.9) | 72.8(7.7) |
| | ADDNEURO | 644 | 0 | 2 | 0 | 0 | 0 | 642 | 186 | 256 | 105(56.5) | 164(64.1) | 76.4(6.6) | 73.0(6.7) |
| | ADGC-ExomeChip | 13969 | 55 | 197 | 12 | 32 | 0 | 13673 | 5250 | 7830 | 3136(59.7) | 4585(58.6) | 79.6(9.0) | 73.0(9.1) |
| | ADNI | 2134 | 63 | 69 | 21 | 30 | 5 | 1945 | 606 | 761 | 260(42.9) | 330(43.4) | 78.5(7.8) | 74.1(7.4) |
| | ADNI3 | 327 | 4 | 12 | 1 | 4 | 0 | 306 | 228 | 24 | 142(62.3) | 10(41.7) | 72.5(6.1) | 72.7(9.5) |
| | CIDR | 3101 | 93 | 2780 | 70 | 0 | 0 | 158 | 1505 | 1530 | 1033(68.6) | 986(64.4) | 74.5(9.4) | 75.5(9.6) |
| | GSK | 1571 | 0 | 1 | 1 | 0 | 0 | 1569 | 773 | 798 | 497(64.3) | 459(57.5) | 73.4(7.9) | 72.5(8.6) |
| | IIDP AA | 1175 | 815 | 359 | 0 | 0 | 0 | 1 | 1001 | 172 | 663(66.2) | 107(62.2) | 83.3(5.3) | 83.6(6.7) |
| | IIDP YOR | 1264 | 1253 | 10 | 0 | 0 | 0 | 1 | 1145 | 104 | 732(63.9) | 79(76.0) | 82.6(5.9) | 77.9(7.2) |
| | LATC | 63 | 13 | 23 | 24 | 0 | 0 | 0 | 15 | 2 | 15(100.0) | 2(100.0) | 77.4(5.4) | 78.0(0.0) |
| | MARS | 708 | 423 | 275 | 1 | 0 | 0 | 1 | 463 | 79 | 392(84.7) | 54(68.4) | 79.6(6.1) | 77.3(7.1) |
| | MAYO | 2413 | 7 | 24 | 2 | 4 | 0 | 2335 | 1225 | 948 | 642(52.4) | 546(57.6) | 75.5(6.5) | 74.0(6.0) |
| | MIRAGE | 1502 | 1 | 28 | 2 | 0 | 0 | 1471 | 738 | 601 | 436(59.1) | 366(60.9) | 72.1(7.3) | 68.8(8.6) |
| | MTC | 542 | 5 | 29 | 12 | 0 | 0 | 496 | 202 | 272 | 130(64.4) | 157(57.7) | 71.7(8.9) | 72.6(9.3) |
| | NIA-LOAD | 5220 | 112 | 642 | 13 | 8 | 0 | 4445 | 2091 | 2351 | 1278(61.1) | 1546(65.8) | 70.6(12.6) | 73.6(7.8) |
| | OHSU | 647 | 3 | 2 | 0 | 1 | 0 | 635 | 379 | 201 | 205(54.1) | 127(63.2) | 85.7(7.5) | 85.0(6.9) |
| | ROSMAP | 2584 | 13 | 50 | 28 | 9 | 0 | 2451 | 1102 | 951 | 795(72.1) | 690(72.6) | 85.4(7.4) | 84.1(6.5) |
| | TARCC | 2718 | 75 | 218 | 821 | 7 | 2 | 1557 | 1124 | 908 | 788(70.1) | 502(55.3) | 70.1(9.8) | 70.1(8.9) |
| | TGEN2 | 1599 | 0 | 9 | 1 | 0 | 1 | 1512 | 573 | 1005 | 255(44.5) | 640(63.7) | 80.8(8.7) | 72.8(8.0) |
| | UM-VU-MSSM | 2462 | 5 | 16 | 0 | 0 | 0 | 2441 | 1195 | 1206 | 724(60.6) | 778(64.5) | 74.1(8.2) | 74.2(7.9) |
| | UPITT | 2440 | 7 | 8 | 1 | 0 | 0 | 2355 | 896 | 1406 | 563(62.8) | 908(64.6) | 75.6(6.2) | 73.2(6.6) |
| | WASHU | 670 | 0 | 0 | 0 | 0 | 0 | 670 | 202 | 429 | 125(61.9) | 239(55.7) | 77.9(8.7) | 74.0(9.6) |
| | WASHU2 | 235 | 10 | 1 | 0 | 0 | 0 | 224 | 116 | 68 | 65(56.0) | 38(55.9) | 73.7(8.6) | 74.0(8.1) |
| | WHICAP | 647 | 0 | 7 | 0 | 0 | 0 | 640 | 554 | 85 | 335(60.5) | 60(70.6) | 82.7(6.7) | 84.1(7.5) |

**Table S5. R145C per cohort, diagnosis and *APOE* genotypes.** DX: diagnosis, N: number of individuals, n: number of R145C allele, Rsq: imputation quality, MAF: R145C minor allele frequency within the considered subset, Ntot: number of individuals within the considered *APOE* genotype.

| | | | | R145C | | | *APOE* ε2ε3 | | | *APOE* ε3ε3 | | | *APOE* ε3ε4 | | |
|---|---|---|---|---|---|---|---|---|---|---|---|---|---|---|---|
| | Cohort | DX | N | n | Rsq | MAF | Ntot | n | MAF | Ntot | n | MAF | Ntot | n | MAF |
| *Discovery* | ADSP WES | AD | 1027 | 40 | Sequenced | 0.019 | 88 | 26 | 0 | 415 | 26 | 0.031 | 386 | 14 | 0.018 |
| | | CN | 2176 | 70 | | 0.016 | 308 | 55 | 0.01 | 1124 | 55 | 0.024 | 588 | 9 | 0.008 |
| | ADSP WGS | AD | 1861 | 77 | Sequenced | 0.021 | 137 | 35 | 0.015 | 730 | 35 | 0.024 | 707 | 38 | 0.027 |
| | | CN | 2781 | 102 | | 0.018 | 401 | 80 | 0.015 | 1498 | 80 | 0.027 | 700 | 10 | 0.007 |
| *Replication* | ACT | AD | 8 | 0 | 0.95 | 0 | 2 | 0 | 0 | 2 | 0 | 0 | 2 | 0 | 0 |
| | | CN | 13 | 2 | | 0.077 | 3 | 2 | 0 | 6 | 2 | 0.167 | 4 | 0 | 0 |
| | ADC1 | AD | 10 | 0 | 0.85 | 0 | 1 | 0 | 0 | 2 | 0 | 0 | 2 | 0 | 0 |
| | | CN | 11 | 0 | | 0 | 5 | 0 | 0 | 0 | 0 | 0 | 6 | 0 | 0 |
| | ADGC-ExomeChip | AD | 24 | 2 | Genotyped | 0.042 | 1 | 1 | 0 | 9 | 1 | 0.056 | 11 | 1 | 0.045 |
| | | CN | 44 | 2 | | 0.023 | 9 | 2 | 0 | 21 | 2 | 0.048 | 13 | 0 | 0 |
| | ADNI | AD | 6 | 1 | 1 | 0.083 | 0 | 1 | 0 | 2 | 1 | 0.25 | 3 | 0 | 0 |
| | | CN | 1 | 0 | | 0 | 0 | 0 | 0 | 0 | 0 | 0 | 0 | 0 | 0 |
| | ADNI3 | AD | 0 | 0 | 1 | 0 | 0 | 0 | 0 | 0 | 0 | 0 | 0 | 0 | 0 |
| | | CN | 5 | 1 | | 0.1 | 0 | 1 | 0 | 2 | 1 | 0.25 | 2 | 0 | 0 |
| | CIDR | AD | 651 | 18 | 0.96 | 0.014 | 35 | 5 | 0 | 238 | 5 | 0.011 | 275 | 13 | 0.024 |
| | | CN | 406 | 9 | | 0.011 | 31 | 7 | 0 | 218 | 7 | 0.016 | 120 | 2 | 0.008 |
| | IIDP AA | AD | 41 | 1 | 0.98 | 0.012 | 5 | 1 | 0 | 17 | 1 | 0.029 | 14 | 0 | 0 |
| | | CN | 642 | 23 | | 0.018 | 116 | 15 | 0.013 | 301 | 15 | 0.025 | 170 | 5 | 0.015 |
| | IIDP YOR | AD | 98 | 5 | 0.99 | 0.026 | 10 | 3 | 0 | 34 | 3 | 0.044 | 39 | 2 | 0.026 |
| | | CN | 1073 | 54 | | 0.025 | 144 | 40 | 0.024 | 502 | 40 | 0.04 | 319 | 7 | 0.011 |
| | LATC | AD | 0 | 0 | 0.95 | 0 | 0 | 0 | 0 | 0 | 0 | 0 | 0 | 0 | 0 |
| | | CN | 9 | 1 | | 0.056 | 2 | 0 | 0 | 1 | 0 | 0 | 5 | 1 | 0.1 |
| | MARS | AD | 44 | 0 | 0.95 | 0 | 7 | 0 | 0 | 20 | 0 | 0 | 11 | 0 | 0 |
| | | CN | 368 | 22 | | 0.03 | 56 | 16 | 0.009 | 181 | 16 | 0.044 | 91 | 5 | 0.027 |
| | NIA-LOAD | AD | 275 | 9 | 0.96 | 0.016 | 14 | 2 | 0.036 | 91 | 2 | 0.011 | 111 | 6 | 0.027 |
| | | CN | 137 | 0 | | 0 | 8 | 0 | 0 | 67 | 0 | 0 | 46 | 0 | 0 |
| | ROSMAP | AD | 11 | 1 | 0.97 | 0.045 | 0 | 1 | 0 | 8 | 1 | 0.062 | 3 | 0 | 0 |
| | | CN | 15 | 1 | | 0.033 | 1 | 1 | 0 | 10 | 1 | 0.05 | 3 | 0 | 0 |
| | TARCC | AD | 32 | 1 | 0.99 | 0.016 | 1 | 0 | 0 | 7 | 0 | 0 | 19 | 1 | 0.026 |
| | | CN | 20 | 3 | | 0.075 | 2 | 2 | 0 | 9 | 2 | 0.111 | 7 | 1 | 0.071 |
| | UM-VU-MSSM | AD | 1 | 0 | 0.94 | 0 | 0 | 0 | 0 | 0 | 0 | 0 | 1 | 0 | 0 |
| | | CN | 0 | 0 | | 0 | 0 | 0 | 0 | 0 | 0 | 0 | 0 | 0 | 0 |

**Table S6. Demographics per cohort after ancestry selection, quality control and duplicates removal.** Dx: diagnosis, AAD: age-at death, AAL: age-at-last-exam, AAE: age-at-exam (and Dx), AAO: age-at-onset.

| | | | | Sex | Age | Age Type | | | |
|---|---|---|---|---|---|---|---|---|---|
| | Cohort | Dx | N | Females (%) | μ(σ) | AAD μ(σ) [%] | AAL μ(σ) [%] | AAE μ(σ) [%] | AAO μ(σ) [%] |
| *Discovery* | ADSP WES | AD | 1027 | 69.0% | 78.4(8.7) | 75.0(-)[0.1%] | - | 79.9(6.9)[4.3%] | 78.3(8.8)[94.1%] |
| | | CN | 2176 | 69.3% | 78.3(7.8) | 80.9(8.4)[1.1%] | 78.3(7.8)[97.7%] | - | - |
| | ADSP WGS | AD | 1861 | 68.5% | 75.3(8.9) | - | - | 74.7(7.5)[0.8%] | 75.3(8.9)[98.7%] |
| | | CN | 2781 | 74.1% | 75.4(8.7) | 82.3(8.8)[3.9%] | 75.1(8.6)[92.4%] | - | - |
| *Replication* | ACT | AD | 8 | 75.0% | 76.1(4.6) | 74.2(3.3)[62.5%] | - | - | 81.0(4.2)[25.0%] |
| | | CN | 13 | 53.8% | 80.4(5.5) | 81.1(5.8)[61.5%] | 79.2(5.4)[38.5%] | - | - |
| | ADC1 | AD | 10 | 80.0% | 71.3(8.1) | - | - | 71.0(-)[10.0%] | 71.3(8.6)[90.0%] |
| | | CN | 11 | 72.7% | 73.8(10.7) | 84.0(-)[9.1%] | 72.8(10.7)[90.9%] | - | - |
| | ADGC-ExomeChip | AD | 24 | 79.2% | 75.0(7.9) | - | - | - | 75.0(7.9)[100.0%] |
| | | CN | 44 | 63.6% | 80.3(6.7) | 80.6(6.7)[54.5%] | 79.9(6.8)[45.5%] | - | - |
| | ADNI | AD | 6 | 83.3% | 73.7(8.9) | - | - | 75.4(8.8)[83.3%] | 65.0(-)[16.7%] |
| | | CN | 1 | 100% | 81.0(-) | - | 81.0(-)[100.0%] | - | - |
| | ADNI3 | AD | 0 | - | - | - | - | - | - |
| | | CN | 5 | 100% | 68.6(6.9) | - | 68.6(6.9)[100.0%] | - | - |
| | CIDR | AD | 651 | 65.6% | 74.0(10.2) | - | - | - | 74.0(10.2)[100.0%] |
| | | CN | 406 | 71.2% | 67.4(8.5) | - | 67.4(8.5)[100.0%] | - | - |
| | IIDP AA | AD | 41 | 75.6% | 87.6(8.8) | - | - | 87.6(8.8)[100.0%] | - |
| | | CN | 642 | 64.0% | 82.9(5.5) | - | 82.9(5.5)[100.0%] | - | - |
| | IIDP YOR | AD | 98 | 74.5% | 78.0(6.8) | - | - | 78.0(6.8)[100.0%] | - |
| | | CN | 1073 | 64.0% | 82.6(5.9) | - | 82.6(5.9)[100.0%] | - | - |
| | LATC | AD | 0 | - | - | - | - | - | - |
| | | CN | 9 | 100% | 73.9(3.3) | - | 73.9(3.3)[100.0%] | - | - |
| | MARS | AD | 44 | 68.2% | 77.4(6.8) | - | - | 77.4(6.8)[100.0%] | - |
| | | CN | 368 | 84.8% | 79.3(6.0) | 80.0(7.7)[17.4%] | 79.2(5.6)[82.6%] | - | - |
| | NIA-LOAD | AD | 275 | 70.9% | 73.9(8.8) | - | - | - | 73.9(8.8)[100.0%] |
| | | CN | 137 | 68.6% | 64.3(10.7) | 78.0(7.9)[3.6%] | 63.7(10.5)[96.4%] | - | - |
| | ROSMAP | AD | 11 | 90.9% | 77.5(4.9) | - | - | 77.1(4.9)[90.9%] | 82.0(-)[9.1%] |
| | | CN | 15 | 93.3% | 79.3(8.4) | 79.8(6.7)[26.7%] | 79.2(9.2)[73.3%] | - | - |
| | TARCC | AD | 32 | 81.2% | 69.9(8.3) | - | - | - | 69.9(8.3)[100.0%] |
| | | CN | 20 | 85.0% | 66.3(10.1) | - | 66.3(10.1)[100.0%] | - | - |
| | UM-VU-MSSM | AD | 1 | 100% | 73.0(-) | - | - | - | 73.0(-)[100.0%] |
| | | CN | 0 | - | - | - | - | - | - |

**Table S7. African ancestry cutoff sensitivity analyses adding age as a covariate.** NB: Age adjustment in the *APOE ε3/ε4* discovery and replication samples is incorrect because controls are older than cases and the model incorrectly infers the age effect on AD risk.

| Sample | | AFR (≥%) | AD Case-Control Regression | | | |
|---|---|---|---|---|---|---|
| | | | N | MAC | OR [95% CI] | P |
| *Discovery* | *APOE ε2ε3* | 15 | 920 | 22 | 0.8[0.28; 2.29] | 0.67 |
| | | 45 | 733 | 20 | 1.05[0.33; 3.29] | 0.94 |
| | | 75 | 463 | 12 | 2.76[0.62; 12.28] | 0.18 |
| | *APOE ε3ε3* | 15 | 3683 | 190 | 1.1[0.8; 1.51] | 0.56 |
| | | 45 | 2431 | 148 | 1.12[0.78; 1.61] | 0.53 |
| | | 75 | 1477 | 101 | 1.19[0.77; 1.84] | 0.43 |
| | *APOE ε3ε4* | 15 | 2341 | 70 | 2.88[1.77; 4.66] | **1.8E-05** |
| | | 45 | 1828 | 65 | 2.94[1.78; 4.85] | **2.4E-05** |
| | | 75 | 1215 | 52 | 3.12[1.78; 5.48] | **7.0E-05** |
| *Replication* | *APOE ε2ε3* | 15 | 452 | 12 | 0.8[0.11; 5.65] | 0.82 |
| | | 45 | 394 | 12 | 0.77[0.11; 5.38] | 0.8 |
| | | 75 | 303 | 9 | 0.31[0.02; 4.73] | 0.4 |
| | *APOE ε3ε3* | 15 | 1748 | 100 | 0.9[0.49; 1.62] | 0.72 |
| | | 45 | 1291 | 91 | 0.9[0.46; 1.76] | 0.77 |
| | | 75 | 923 | 77 | 0.64[0.3; 1.37] | 0.25 |
| | *APOE ε3ε4* | 15 | 1277 | 44 | 2.2[1.04; 4.65] | **0.04** |
| | | 45 | 900 | 31 | 1.96[0.75; 5.09] | **0.17** |
| | | 75 | 632 | 22 | 1.47[0.45; 4.82] | **0.53** |
| ***Meta-analysis*** | *APOE ε2ε3* | 15 | 1372 | 34 | 0.8[0.32; 2.02] | 0.63 |
| | | 45 | 1127 | 32 | 0.97[0.36; 2.6] | 0.95 |
| | | 75 | 766 | 21 | 1.66[0.45; 6.16] | 0.44 |
| | *APOE ε3ε3* | 15 | 5431 | 290 | 1.05[0.79; 1.39] | 0.74 |
| | | 45 | 3722 | 239 | 1.07[0.78; 1.47] | 0.68 |
| | | 75 | 2400 | 178 | 1.02[0.7; 1.49] | 0.91 |
| | *APOE ε3ε4* | 15 | 3618 | 114 | 2.66[1.77; 3.99] | **2.3E-06** |
| | | 45 | 2728 | 96 | 2.69[1.73; 4.2] | **1.2E-05** |
| | | 75 | 1847 | 74 | 2.72[1.64; 4.52] | **1.1E-04** |

**Table S8. African ancestry cutoff sensitivity analyses.** Three representative thresholds are shown: 15%, 45%, and 75% corresponding respectively to the threshold use in the main analysis (15%), a first generation admix individual (45%), and the traditional cut off for super ancestry assignment in SNPWeight (75%). Overall, the results are very similar for the three cutoffs and the main finding in the *APOE* ε3/ε4 remains unchanged. Some analyses are more significant or show larger effect size at 45% or 75% cutoffs or other intermediate values (data not shown) than at 15% cutoff.

| Sample | | AFR (≥%) | AD Case-Control Regression N | MAC | OR [95% CI] | P | AD Age-at-onset Regression N | MAC | β [95% CI] | P | Competing Risk Regression N | MAC | HR [95% CI] | P |
|---|---|---|---|---|---|---|---|---|---|---|---|---|---|---|
| *Discovery* | *APOE ε2ε3* | 15 | 934 | 22 | 0.73 [0.26; 2.04] | 0.55 | 222 | 4 | -6.96 [-15.56; 1.64] | 0.11 | 918 | 22 | 1.14 [0.35; 3.69] | 0.82 |
| | | 45 | 738 | 20 | 0.9 [0.3; 2.66] | 0.84 | 163 | 4 | -6.84 [-15.17; 1.5] | 0.11 | 731 | 20 | 1.8 [0.57; 5.64] | 0.31 |
| | | 75 | 465 | 12 | 2.2 [0.53; 9.13] | 0.28 | 95 | 4 | -6.68 [-15.43; 2.06] | 0.13 | 461 | 12 | 3.5 [1.02; 11.96] | 0.05 |
| | *APOE ε3ε3* | 15 | 3767 | 196 | 1.06 [0.78; 1.46] | 0.71 | 1108 | 58 | -1.68 [-3.87; 0.5] | 0.13 | 3646 | 187 | 1.09 [0.82; 1.45] | 0.57 |
| | | 45 | 2449 | 149 | 1.12 [0.78; 1.61] | 0.54 | 676 | 44 | -0.01 [-2.63; 2.61] | 0.99 | 2396 | 145 | 1.02 [0.75; 1.38] | 0.91 |
| | | 75 | 1487 | 102 | 1.19 [0.77; 1.84] | 0.43 | 400 | 30 | 0.38 [-2.84; 3.6] | 0.82 | 1459 | 98 | 0.97 [0.67; 1.4] | 0.86 |
| | *APOE ε3ε4* | 15 | 2381 | 71 | 3.01 [1.87; 4.85] | **6.0E-06** | 1063 | 51 | -5.87 [-8.35; -3.4] | **3.4E-06** | 2315 | 70 | 2.66 [1.86; 3.8] | **8.5E-08** |
| | | 45 | 1845 | 66 | 3.17 [1.93; 5.2] | **4.7E-06** | 833 | 48 | -5.48 [-8.04; -2.92] | **2.7E-05** | 1806 | 65 | 2.65 [1.86; 3.79] | **8.6E-08** |
| | | 75 | 1227 | 53 | 3.4 [1.95; 5.9] | **1.5E-05** | 552 | 39 | -4.98 [-7.76; -2.19] | **4.6E-04** | 1201 | 52 | 2.59 [1.76; 3.82] | **1.5E-06** |
| *Replication* | *APOE ε2ε3* | 15 | 453 | 12 | 0.78 [0.11; 5.35] | 0.8 | 53 | 1 | -18.42 [-39.23; 2.38] | 0.08 | 430 | 12 | 3.05 [0.33; 28.09] | 0.32 |
| | | 45 | 395 | 12 | 0.79 [0.11; 5.5] | 0.81 | 23 | 1 | -24.18 [-46.48; -1.89] | 0.03 | 372 | 12 | 2.17 [0.14; 34.15] | 0.58 |
| | | 75 | 304 | 9 | 0.32 [0.02; 4.92] | 0.41 | - | - | - | - | - | - | - | - |
| | *APOE ε3ε3* | 15 | 1748 | 100 | 0.85 [0.48; 1.53] | 0.6 | 347 | 8 | -1.36 [-8.29; 5.58] | 0.7 | 1656 | 94 | 0.8 [0.41; 1.57] | 0.51 |
| | | 45 | 1291 | 91 | 0.93 [0.47; 1.81] | 0.83 | 108 | 4 | -7.36 [-17.38; 2.66] | 0.15 | 1206 | 85 | 0.98 [0.35; 2.79] | 0.98 |
| | | 75 | 923 | 77 | 0.78 [0.36; 1.69] | 0.53 | 19 | 2 | -12.1 [-28.68; 4.47] | 0.15 | 857 | 73 | 1.39 [0.34; 5.61] | 0.64 |
| | *APOE ε3ε4* | 15 | 1277 | 44 | 2.2 [1.04; 4.65] | **0.04** | 421 | 21 | -5.23 [-9.58; -0.87] | **0.02** | 1195 | 42 | 2.0 [1.19; 3.35] | **8.7E-03** |
| | | 45 | 900 | 31 | 2.48 [0.95; 6.5] | **0.06** | 170 | 10 | -5.5 [-11.34; 0.33] | **0.06** | 826 | 29 | 2.51 [1.31; 4.81] | **5.5E-03** |
| | | 75 | 632 | 22 | 2.53 [0.75; 8.5] | **0.13** | 45 | 4 | -6.62 [-15.89; 2.66] | **0.16** | 576 | 20 | 4.82 [1.67; 13.9] | **3.6E-03** |
| ***Meta-analysis*** | *APOE ε2ε3* | 15 | 1387 | 34 | 0.74 [0.3; 1.84] | 0.52 | 275 | 5 | -8.63 [-16.58; -0.69] | 0.03 | 1348 | 34 | 1.42 [0.5; 3.99] | 0.51 |
| | | 45 | 1133 | 32 | 0.87 [0.34; 2.24] | 0.77 | 186 | 5 | -8.96 [-16.77; -1.15] | 0.02 | 1103 | 32 | 1.85 [0.64; 5.31] | 0.25 |
| | | 75 | 769 | 21 | 1.45 [0.41; 5.15] | 0.56 | 95 | 4 | -6.68 [-15.43; 2.06] | 0.13 | 461 | 12 | 3.5 [1.02; 11.96] | 0.05 |
| | *APOE ε3ε3* | 15 | 5515 | 296 | 1.01 [0.77; 1.34] | 0.94 | 1455 | 66 | -1.65 [-3.74; 0.43] | 0.12 | 5302 | 281 | 1.04 [0.8; 1.35] | 0.79 |
| | | 45 | 3740 | 240 | 1.07 [0.78; 1.47] | 0.66 | 784 | 48 | -0.48 [-3.02; 2.05] | 0.71 | 3602 | 230 | 1.01 [0.75; 1.36] | 0.92 |
| | | 75 | 2410 | 179 | 1.08 [0.74; 1.57] | 0.7 | 419 | 32 | -0.07 [-3.23; 3.09] | 0.96 | 2316 | 171 | 0.99 [0.69; 1.42] | 0.96 |
| | *APOE ε3ε4* | 15 | 3658 | 115 | 2.75 [1.84; 4.11] | **8.3E-07** | 1484 | 72 | -5.72 [-7.87; -3.56] | **2.0E-07** | 3510 | 112 | 2.42 [1.81; 3.25] | **3.7E-09** |
| | | 45 | 2745 | 97 | 3.01 [1.94; 4.68] | **8.9E-07** | 1003 | 58 | -5.49 [-7.83; -3.14] | **4.4E-06** | 2632 | 94 | 2.62 [1.92; 3.58] | **1.6E-09** |
| | | 75 | 1859 | 75 | 3.23 [1.95; 5.34] | **5.0E-06** | 597 | 43 | -5.11 [-7.78; -2.45] | **1.7E-04** | 1777 | 72 | 2.79 [1.94; 4.02] | **3.5E-08** |

**Table S9. African ancestry cutoff sensitivity analyses restricted to individuals with AFR local ancestry at both of their *APOE* haplotypes.**

| | | AD Case-Control Regression | | | | AD Age-at-onset Regression | | | | Competing Risk Regression | | | |
|---|---|---|---|---|---|---|---|---|---|---|---|---|---|
| Sample | AFR (≥%) | N | MAC | OR [95% CI] | P | N | MAC | β [95% CI] | P | N | MAC | HR [95% CI] | P |
| *APOE* ε3ε4 | 15 | 978 | 44 | 3.10 [1.68; 5.7] | 2.8E-04 | 434 | 32 | -5.11 [-8.19; -2.03] | 1.1E-03 | 955 | 44 | 2.50 [1.62; 3.86] | 3.7E-05 |
| | 45 | 917 | 44 | 3.16 [1.72; 5.82] | 2.2E-04 | 409 | 32 | -5.10 [-8.24; -1.96] | 1.4E-03 | 895 | 44 | 2.52 [1.63; 3.88] | 2.9E-05 |
| | 75 | 719 | 36 | 3.1 [1.57; 6.13] | 1.1E-03 | 321 | 26 | -4.78 [-8.25; -1.31] | 7.0E-03 | 703 | 36 | 2.37 [1.44; 3.9] | 7.2E-04 |

#### Table S10. Mega-analysis of the discovery and replication samples.

| Sample | | AFR (≥%) | AD Case-Control Regression N | MAC | OR [95% CI] | P | AD Age-at-onset Regression N | MAC | β [95% CI] | P | Competing Risk Regression N | MAC | HR [95% CI] | P |
|---|---|---|---|---|---|---|---|---|---|---|---|---|---|---|
| ***Mega-analysis*** | *APOE* ε2ε3 | 15 | 1387 | 34 | 0.76 [0.31; 1.87] | 0.55 | 275 | 5 | -9.07 [-17.08; -1.06] | 0.03 | 1226 | 32 | 1.16 [0.4; 3.35] | 0.78 |
| | | 45 | 1133 | 32 | 0.87 [0.34; 2.2] | 0.76 | 186 | 5 | -8.97 [-16.69; -1.26] | 0.02 | 1016 | 30 | 1.75 [0.65; 4.69] | 0.27 |
| | | 75 | 769 | 21 | 1.51 [0.43; 5.26] | 0.52 | 100 | 4 | -7.21 [-16.02; 1.6] | 0.11 | 411 | 12 | 3.27 [1.11; 9.62] | 0.03 |
| | *APOE* ε3ε3 | 15 | 5515 | 296 | 0.98 [0.75; 1.28] | 0.86 | 1455 | 66 | -1.36 [-3.5; 0.77] | 0.21 | 4785 | 260 | 1.08 [0.84; 1.39] | 0.55 |
| | | 45 | 3740 | 240 | 1.05 [0.77; 1.44] | 0.74 | 784 | 48 | -0.32 [-2.88; 2.24] | 0.81 | 3288 | 213 | 1.04 [0.78; 1.39] | 0.78 |
| | | 75 | 2410 | 179 | 1.04 [0.72; 1.49] | 0.84 | 419 | 32 | -0.06 [-3.2; 3.09] | 0.97 | 2159 | 161 | 1.04 [0.73; 1.46] | 0.84 |
| | *APOE* ε3ε4 | 15 | **3658** | **115** | **2.93 [1.99; 4.31]** | **4.8E-08** | **1484** | **72** | **-5.86 [-8.05; -3.66]** | **1.7E-07** | **3233** | **106** | **2.67 [2.01; 3.54]** | **1.3E-11** |
| | | 45 | **2745** | **97** | **3.02 [1.97; 4.61]** | **3.6E-07** | **1003** | **58** | **-5.7 [-8.06; -3.33]** | **2.4E-06** | **2451** | **91** | **2.73 [1.99; 3.74]** | **4.2E-10** |
| | | 75 | **1859** | **75** | **3.25 [1.99; 5.3]** | **2.4E-06** | **597** | **43** | **-5.35 [-8.01; -2.69]** | **8.0E-05** | **1679** | **71** | **2.73 [1.9; 3.93]** | **5.5E-08** |

#### Table S11. Primary and secondary analyses considering a standard model (non-stratified by *APOE* genotype) and adjusting for ε2 and ε4 dosages.

| | | AD Case-Control Regression | | | | AD Age-at-onset Regression | | | | Competing Risk Regression | | | |
|---|---|---|---|---|---|---|---|---|---|---|---|---|---|
| Sample | AFR (≥%) | N | MAC | OR [95% CI] | P | N | MAC | β [95% CI] | P | N | MAC | HR [95% CI] | P |
| *Discovery* | 15 | 7845 | 289 | 1.40 [1.08; 1.8] | 0.01 | 2802 | 113 | -3.55 [-5.15; -1.95] | 1.4E-05 | 7612 | 279 | 1.49 [1.21; 1.84] | 1.7E-04 |
| | 45 | 5684 | 235 | 1.56 [1.18; 2.07] | 2.0E-03 | 2030 | 96 | -3.01 [-4.8; -1.21] | 1.0E-03 | 5567 | 230 | 1.57 [1.26; 1.96] | 4.8E-05 |
| | 75 | 3633 | 167 | 1.8 [1.29; 2.51] | 5.1E-04 | 1298 | 73 | -2.81 [-4.82; -0.79] | 6.3E-03 | 3567 | 162 | 1.61 [1.25; 2.07] | 2.3E-04 |
| *Replication* | 15 | 3945 | 156 | 1.23 [0.78; 1.94] | 0.38 | 996 | 30 | -4.42 [-7.98; -0.86] | 0.01 | 3716 | 148 | 1.37 [0.95; 1.97] | 0.09 |
| | 45 | 2928 | 134 | 1.28 [0.75; 2.17] | 0.37 | 378 | 15 | -6.80 [-11.76; -1.83] | 7.3E-03 | 2716 | 126 | 1.60 [0.95; 2.68] | 0.08 |
| | 75 | 2086 | 108 | 1.03 [0.55; 1.94] | 0.93 | 90 | 6 | -8.54 [-15.72; -1.35] | 0.02 | 1918 | 102 | 2.64 [1.14; 6.11] | 0.02 |
| *Meta-analysis* | 15 | 11790 | 445 | 1.36 [1.08; 1.69] | 7.5E-03 | 3798 | 143 | -3.70 [-5.16; -2.24] | 7.0E-07 | 11328 | 427 | 1.46 [1.22; 1.75] | 4.1E-05 |
| | 45 | 8612 | 369 | 1.49 [1.16; 1.91] | 1.6E-03 | 2408 | 111 | -3.44 [-5.13; -1.76] | 6.3E-05 | 8283 | 356 | 1.58 [1.29; 1.93] | 9.3E-06 |
| | 75 | 5719 | 275 | 1.6 [1.19; 2.14] | 1.8E-03 | 1388 | 79 | -3.23 [-5.17; -1.28] | 1.1E-03 | 5485 | 264 | 1.68 [1.32; 2.14] | 2.9E-05 |

#### Table S12. Interaction between R145C*ε4 for association with AD risk, considering a standard model (non-stratified by *APOE* genotype) and adjusting for ε2 and ε4 dosages.

| | | AD Case-Control Regression | | | |
|---|---|---|---|---|---|
| Sample | AFR (≥%) | N | MAC | OR – R145C*ε4 [95% CI] | P |
| *Discovery* | 15 | 7845 | 289 | 2.73 [1.57; 4.75] | 3.6E-04 |
| | 45 | 5684 | 235 | 2.58 [1.43; 4.67] | 1.7E-03 |
| | 75 | 3633 | 167 | 2.38 [1.20; 4.70] | 0.01 |
| *Replication* | 15 | 3945 | 156 | 2.51 [1.08; 5.85] | 0.03 |
| | 45 | 2928 | 134 | 2.71 [0.91; 8.05] | 0.07 |
| | 75 | 2086 | 108 | 3.16 [0.84; 11.87] | 0.09 |
| *Meta-analysis* | 15 | 11790 | 445 | 2.66 [1.68; 4.23] | 3.4E-05 |
| | 45 | 8612 | 369 | 2.61 [1.55; 4.40] | 3.1E-04 |
| | 75 | 5719 | 275 | 2.52 [1.38; 4.63] | 2.8E-03 |

**Table S13.** ***APOE*** **variants genotype call in the data released by NIAGADS.** First set of columns corresponds to number of individuals missing either of the three SNPs, the second set of columns to individuals missing any pair of three SNPs, and third set corresponds to individuals missing all three SNPs.

| Cohort N | Missing genotypes – N (%) | | | | | | |
|---|---|---|---|---|---|---|---|
| | **rs429358** | **rs769455** | **rs7412** | **rs429358 & rs769455** | **rs429358 & rs7412** | **rs769455 & rs7412** | **rs429358 & rs769455 & rs7412** |
| **ADSP WES** 20,504 | 760 (3.7%) | 3,730 (18.2%) | 6,661 (32.5%) | 667 (3.3%) | 701 (3.4%) | 3,607 (17.6%) | 666 (3.2%) |
| **ADSP WGS** 16,906 | 330 (2.0%) | 499 (3.0%) | 481 (2.8%) | 278 (1.6%) | 263 (1.6%) | 442 (2.6%) | 256 (1.5%) |

**Table S14. African ancestry cutoff sensitivity analyses only including individuals directly genotyped in the ADSP WES data distributed by NIAGADS.** Note that compare to **Table S8**, solely the discovery sample changed by removing the 980 individuals who had any of the 3 APOE missenses variant missing in the NIAGADS call of ADSP WES. (**Figure S3**)._As in **Table S8**, three representative thresholds are shown: 15%, 45%, and 75% corresponding respectively to the threshold use in the main analysis (15%), a first generation admix individual (45%), and the traditional cut off for super ancestry assignment in SNPWeight (75%). Overall, the results are very similar for the three cutoffs and the main finding in the *APOE* ε3/ε4 remains unchanged. Some analyses are more significant or show larger effect size at 45% or 75% cutoffs or other intermediate values (data not shown) than at 15% cutoff.

| | | | AD Case-Control Regression | | | | AD Age-at-onset Regression | | | | Competing Risk Regression | | | |
|---|---|---|---|---|---|---|---|---|---|---|---|---|---|---|
| Sample | | AFR (≥%) | N | MAC | OR [95% CI] | P | N | MAC | β [95% CI] | P | N | MAC | HR [95% CI] | P |
| *Discovery* | *APOE ε2ε3* | 15 | 812 | 20 | 0.84 [0.28; 2.46] | 0.75 | 196 | 4 | -6.35 [-14.72; 2.01] | 0.14 | 796 | 20 | 1.4 [0.42; 4.68] | 0.58 |
| | | 45 | 651 | 18 | 1.02 [0.33; 3.22] | 0.97 | 147 | 4 | -6.26 [-14.54; 2.01] | 0.14 | 644 | 18 | 2.46 [0.79; 7.67] | 0.12 |
| | | 75 | 415 | 12 | 2.17 [0.52; 9.06] | 0.29 | 87 | 4 | -6.31 [-15.03; 2.41] | 0.16 | 411 | 12 | 3.29 [0.99; 11.01] | 0.05 |
| | *APOE ε3ε3* | 15 | 3247 | 175 | 1.05 [0.75; 1.46] | 0.79 | 983 | 53 | -1.56 [-3.87; 0.75] | 0.19 | 3129 | 166 | 1.05 [0.77; 1.42] | 0.76 |
| | | 45 | 2133 | 132 | 1.19 [0.81; 1.73] | 0.38 | 616 | 42 | -0.31 [-2.97; 2.34] | 0.82 | 2082 | 128 | 1.04 [0.75; 1.44] | 0.81 |
| | | 75 | 1328 | 92 | 1.26 [0.8; 1.99] | 0.31 | 374 | 29 | 0.28 [-2.99; 3.54] | 0.87 | 1302 | 88 | 0.99 [0.68; 1.44] | 0.95 |
| | *APOE ε3ε4* | 15 | 2103 | 65 | 3.1 [1.88; 5.1] | **9.2E-06** | 989 | 49 | -5.65 [-8.2; -3.09] | **1.4E-05** | 2038 | 64 | 2.52 [1.74; 3.65] | **1.1E-06** |
| | | 45 | 1663 | 63 | 3.23 [1.95; 5.36] | **5.4E-06** | 798 | 48 | -5.24 [-7.8; -2.68] | **6.1E-05** | 1625 | 62 | 2.55 [1.78; 3.66] | **3.8E-07** |
| | | 75 | 1128 | 52 | 3.17 [1.82; 5.55] | **5.0E-05** | 540 | 39 | -4.82 [-7.6; -2.04] | **6.9E-04** | 1103 | 51 | 2.39 [1.62; 3.54] | **1.3E-05** |
| *Replication* | *APOE ε2ε3* | 15 | 453 | 12 | 0.78 [0.11; 5.35] | 0.8 | 53 | 1 | -18.42 [-39.23; 2.38] | 0.08 | 430 | 12 | 3.05 [0.33; 28.09] | 0.32 |
| | | 45 | 395 | 12 | 0.79 [0.11; 5.5] | 0.81 | 23 | 1 | -24.18 [-46.48; -1.89] | 0.03 | 372 | 12 | 2.17 [0.14; 34.15] | 0.58 |
| | | 75 | 304 | 9 | 0.32 [0.02; 4.92] | 0.41 | - | - | - | - | - | - | - | - |
| | *APOE ε3ε3* | 15 | 1748 | 100 | 0.85 [0.48; 1.53] | 0.6 | 347 | 8 | -1.36 [-8.29; 5.58] | 0.7 | 1656 | 94 | 0.8 [0.41; 1.57] | 0.51 |
| | | 45 | 1291 | 91 | 0.93 [0.47; 1.81] | 0.83 | 108 | 4 | -7.36 [-17.38; 2.66] | 0.15 | 1206 | 85 | 0.98 [0.35; 2.79] | 0.98 |
| | | 75 | 923 | 77 | 0.78 [0.36; 1.69] | 0.53 | 19 | 2 | -12.1 [-28.68; 4.47] | 0.15 | 857 | 73 | 1.39 [0.34; 5.61] | 0.64 |
| | *APOE ε3ε4* | 15 | 1277 | 44 | 2.2 [1.04; 4.65] | **0.04** | 421 | 21 | -5.23 [-9.58; -0.87] | **0.02** | 1195 | 42 | 2.0 [1.19; 3.35] | **8.7E-03** |
| | | 45 | 900 | 31 | 2.48 [0.95; 6.5] | **0.06** | 170 | 10 | -5.5 [-11.34; 0.33] | **0.06** | 826 | 29 | 2.51 [1.31; 4.81] | **5.5E-03** |
| | | 75 | 632 | 22 | 2.53 [0.75; 8.5] | **0.13** | 45 | 4 | -6.62 [-15.89; 2.66] | **0.16** | 576 | 20 | 4.82 [1.67; 13.9] | **3.6E-03** |
| ***Meta-analysis*** | *APOE ε2ε3* | 15 | 1265 | 32 | 0.82 [0.32; 2.11] | 0.68 | 249 | 5 | -8.03 [-15.79; -0.27] | 0.04 | 1226 | 32 | 1.67 [0.58; 4.83] | 0.34 |
| | | 45 | 1046 | 30 | 0.96 [0.36; 2.57] | 0.93 | 170 | 5 | -8.43 [-16.19; -0.67] | 0.03 | 1016 | 30 | 2.42 [0.84; 6.91] | 0.1 |
| | | 75 | 719 | 21 | 1.44 [0.41; 5.11] | 0.57 | 87 | 4 | -6.31 [-15.03; 2.41] | 0.16 | 411 | 12 | 3.29 [0.99; 11.01] | 0.05 |
| | *APOE ε3ε3* | 15 | 4995 | 275 | 1.0 [0.75; 1.33] | 0.98 | 1330 | 61 | -1.54 [-3.73; 0.65] | 0.17 | 4785 | 260 | 1.0 [0.76; 1.32] | 0.99 |
| | | 45 | 3424 | 223 | 1.12 [0.8; 1.55] | 0.51 | 724 | 46 | -0.78 [-3.35; 1.79] | 0.55 | 3288 | 213 | 1.04 [0.76; 1.41] | 0.82 |
| | | 75 | 2251 | 169 | 1.12 [0.75; 1.65] | 0.58 | 393 | 31 | -0.19 [-3.4; 3.02] | 0.91 | 2159 | 161 | 1.01 [0.7; 1.46] | 0.95 |
| | *APOE ε3ε4* | 15 | 3380 | 109 | 2.79 [1.84; 4.22] | **1.3E-06** | 1410 | 70 | -5.54 [-7.74; -3.34] | **8.2E-07** | 3233 | 106 | 2.33 [1.72; 3.14] | **3.9E-08** |
| | | 45 | 2563 | 94 | 3.05 [1.95; 4.78] | **1.0E-06** | 968 | 58 | -5.28 [-7.63; -2.94] | **1.0E-05** | 2451 | 91 | 2.54 [1.85; 3.48] | **7.2E-09** |
| | | 75 | 1760 | 74 | 3.05 [1.84; 5.06] | **1.6E-05** | 585 | 43 | -4.96 [-7.63; -2.3] | **2.6E-04** | 1679 | 71 | 2.6 [1.8; 3.75] | **3.4E-07** |
